# Spatial proteo-transcriptomic profiling reveals the molecular landscape of borderline ovarian tumors and their invasive progression

**DOI:** 10.1101/2023.11.13.23298409

**Authors:** Lisa Schweizer, Rahul Krishnan, Aasa Shimizu, Andreas Metousis, Hilary Kenny, Rachelle Mendoza, Thierry M. Nordmann, Sarah Rauch, Lucy Kelliher, Janna Heide, Florian A. Rosenberger, Agnes Bilecz, Sanaa Nakad Borrego, Maximillian T. Strauss, Marvin Thielert, Edwin Rodriguez, Johannes B. Müller-Reif, Mengjie Chen, S. Diane Yamada, Andreas Mund, Ricardo R. Lastra, Matthias Mann, Ernst Lengyel

**Affiliations:** Proteomics and Signal Transduction, Max Planck Institute of Biochemistry, Martinsried, Germany; Department of Obstetrics and Gynecology/Section of Gynecologic Oncology, University of Chicago, Chicago, IL, USA; Proteomics Program, Novo Nordisk Foundation Center for Protein Research, Faculty of Health and Medical Sciences, University of Copenhagen, Copenhagen, Denmark; Department of Pathology, The University of Chicago, Chicago, IL, USA; Medicine/Section of Genetic Medicine, The University of Chicago, Chicago, IL, USA

## Abstract

Serous borderline tumors (SBT) are epithelial neoplastic lesions of the ovaries that commonly have a good prognosis. In 10-15% of cases, however, SBT will recur as low-grade serous cancer (LGSC), which is deeply invasive and responds poorly to current standard chemotherapy^1,2,3^. While genetic alterations suggest a common origin, the transition from SBT to LGSC remains poorly understood^4^. Here, we integrate spatial proteomics^5^ with spatial transcriptomics to elucidate the evolution from SBT to LGSC and its corresponding metastasis at the molecular level in both the stroma and the tumor. We show that the transition of SBT to LGSC occurs in the epithelial compartment through an intermediary stage with micropapillary features (SBT-MP), which involves a gradual increase in MAPK signaling. A distinct subset of proteins and transcripts was associated with the transition to invasive tumor growth, including the neuronal splicing factor NOVA2, which was limited to expression in LGSC and its corresponding metastasis. An integrative pathway analysis exposed aberrant molecular signaling of tumor cells supported by alterations in angiogenesis and inflammation in the tumor microenvironment. Integration of spatial transcriptomics and proteomics followed by knockdown of the most altered genes or pharmaceutical inhibition of the most relevant targets confirmed their functional significance in regulating key features of invasiveness. Combining cell-type resolved spatial proteomics and transcriptomics allowed us to elucidate the sequence of tumorigenesis from SBT to LGSC. The approach presented here is a blueprint to systematically elucidate mechanisms of tumorigenesis and find novel treatment strategies.

## Introduction

Serous ovarian tumors are the most common ovarian cancers. They are characterized as either high-grade (HGSC) or low-grade (LGSC), the latter displaying only mild atypia and few mitotic figures (Fig. 1a). Patients with LGSC tend to be younger than patients with HGSC and have slower growing, widely invasive tumors surrounded by dense fibrotic stroma, making complete surgical removal challenging. If the tumor cannot be entirely removed, metastatic LGSC (LGSC-Met) has an indolent clinical course with late recurrences and a low chance of cure. Although it is a different disease entity, LGSC is treated with the same standard platinum/taxane chemotherapy as HGSC. In LGSC, however, this treatment generally results in minimal clinical response and continued slow progression^1^.

**Fig. 1.**
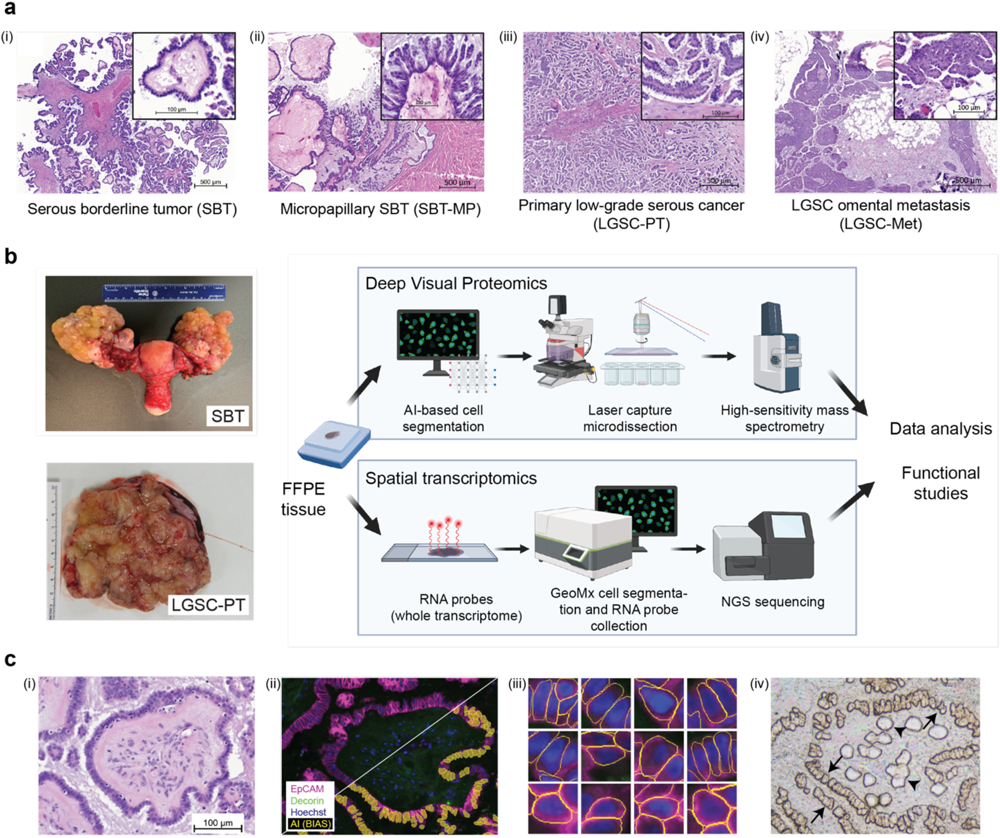
Characterization of serous borderline and low-grade ovarian cancer. **a)** Representative H&E images of the putative transformation sequence from (i) serous borderline tumor (SBT, not invasive) via (ii) micropapillary lesions (SBT-MP) to (iii) low-grade serous cancer-primary tumor (LGSC-PT), and (iv) metastasis (LGSC-Met). The papillary architecture with hierarchical branching pattern is characteristic of SBT. **b)** Gross pathology of a bilateral borderline tumor and low-grade serous cancer. SBT often present as bilateral adnexal tumors (left). Experimental strategy, bioinformatics and functional studies (right). **c)** AI-based cell recognition and laser-based dissection in DVP. (i) H&E of an SBT. (ii, iii) Immunofluorescence. Staining for malignant epithelial cells (EpCAM, purple) and stromal (decorin, green) as well as artificial intelligence (AI)-based recognition of tumor cells (yellow – below diagonal line) using the BIAS software. (iii) Morphology of single epithelial cells as recognized by artificial intelligence. (iv) Brightfield image of the same sample in i and ii showing the tissue after single cell extraction. Microdissected epithelial cells (arrows) and stroma (arrowheads). Serous borderline tumor (SBT), micropapillary SBT (MP-SBT), low-grade serous cancer primary tumor (LGSC-PT) and corresponding metastases (LGSC-Met).

Mutational profiling has suggested that serous borderline tumors (SBT) are precursor lesions of LGSC^4^. SBT are neoplasms of epithelial origin, which mostly are unilateral without any stromal invasion (Fig. 1a, b). Patients with SBT usually have an excellent prognosis after operative tumor removal, but 10-15% will recur with an LGSC, which has very low response rates to current treatment strategies^2,3^. In contrast to HGSC, SBT and LGSC have wild-type p53, few DNA copy-number changes, and a low rate of somatic mutations. Both SBT and LGSC have mutually exclusive mutations in *BRAF*, *KRAS*, or *ERBB2*^6–8^, all upstream regulators of the mitogen-activated protein kinase (MAPK), known to drive cell proliferation. A role for the MAPK pathway and estrogen receptor signaling has been identified in SBT and LGSC, but otherwise, little is known about their molecular landscape. This knowledge is a prerequisite to the identification of novel treatment options.

Here we set out to study the malignant transformation of SBT through putative intermediary steps (micropapillary SBT) to invasiveness in LGSC with spatial omics technologies. Deep Visual Proteomics (DVP) integrates high-content imaging, artificial intelligence (AI)-based cell recognition, laser microdissection, and mass spectrometry to preserve spatial information while quantifying the proteome in an unbiased manner^5^. We combined DVP with probe-based spatial transcriptomics to discover novel pathway alterations in LGSC and to find more effective treatments from the changes in protein and transcript expression during the transition from SBT to LGSC. The transcripts and proteins discovered with this approach included NOVA2, which is not normally expressed in the reproductive system but appears during the LGSC transition. We confirmed the relevance of the candidates identified by spatial omics by targeted functional screens in which a knock-down of the respective genes reduced invasion and migration.

## Results

### Spatial multi-omics of the transition of borderline tumor to low-grade serous cancer

Mutational profiling and histologic observations suggest a progression from serous borderline tumors (SBT) to micropapillary SBT (SBT-MP) and ultimately to low-grade serous carcinoma (LGSC), which may metastasize if not diagnosed and treated early^9^ (Fig. 1a). Histologically, this hypothetical transition is characterized by irregularly contoured papillae with a hierarchical branching pattern in SBT that may progress to a micropapillary tissue architecture^10^, with mild cytologic atypia, signifying a higher risk of progression to LGSC^11^. In contrast to these non-invasive phenotypes, once cells evolve to LGSC, they destructively invade into the stroma and frequently metastasize to the omentum^12^.

We employed spatial proteomics (DVP), and transcriptomics (GeoMX) on a cohort of patients with SBT, SBT-MP, primary LGSC-PT, and corresponding omental metastasis (LGSC-Met) (Supplementary Table 1) for compartment-resolved characterization of different cell types (Fig. 1b). Using specific markers for epithelium (EPCAM) and stroma (Decorin), we stained all tissues with immunofluorescence and recorded high-content images. For DVP, cell populations were segmented on the images using the nucleAIzer AI algorithm^13^ integrated in Biology Image Analysis Software (BIAS), and laser microdissected for ultra-high sensitivity mass spectrometry (MS) data acquisition (Fig. 1c)^14^. DVP detected and quantified a median of 5,456 different proteins in the epithelium and 3,919 in the stroma (Supplementary Table 2), with little inter-and intra-specimen variability and excellent data completeness in all anatomic regions (Extended Data 1 a-f). A unique advantage of DVP is that it records and preserves the morphological characteristics of each cell within the tissue architecture. This is illustrated by more uniform cell sizes (Extended Data 1 g) and a longer distance to the closest cellular neighbor (Extended Data 1h) in LGSC-PT as compared to SBT, the latter being indicative of stromal invasion. All proteins and transcripts highlighted in the following sections are described in Supplementary Table 3.

### Progression of borderline tumors to metastatic low-grade serous carcinoma revealed by DVP

Principal Component Analysis (PCA) of our spatial proteomics results in the epithelial compartment clustered and ordered all four histologies (SBT, SBT-MP, LGSC-PT, LGSC-Met) sequentially. The analysis placed micropapillary growth as an intermediary stage between non-invasive serous borderline and invasive low-grade cancer (Fig. 2a). The LGSC and subsequent metastasis clustered closely, indicating that cells of the primary tumor and the metastasis were very similar after the transition to invasiveness. Linear regression analysis of the four histological stages showed an increase or decrease in 195 proteins in the invasive phenotypes; this panel of proteins represents an interesting subset representing the entire transition from SBT to LGSC and corresponding metastasis (adj. p-value <= 0.05, logFC > 1.5, Extended Data 2a, b, Supplementary Table 4).

**Fig. 2.**
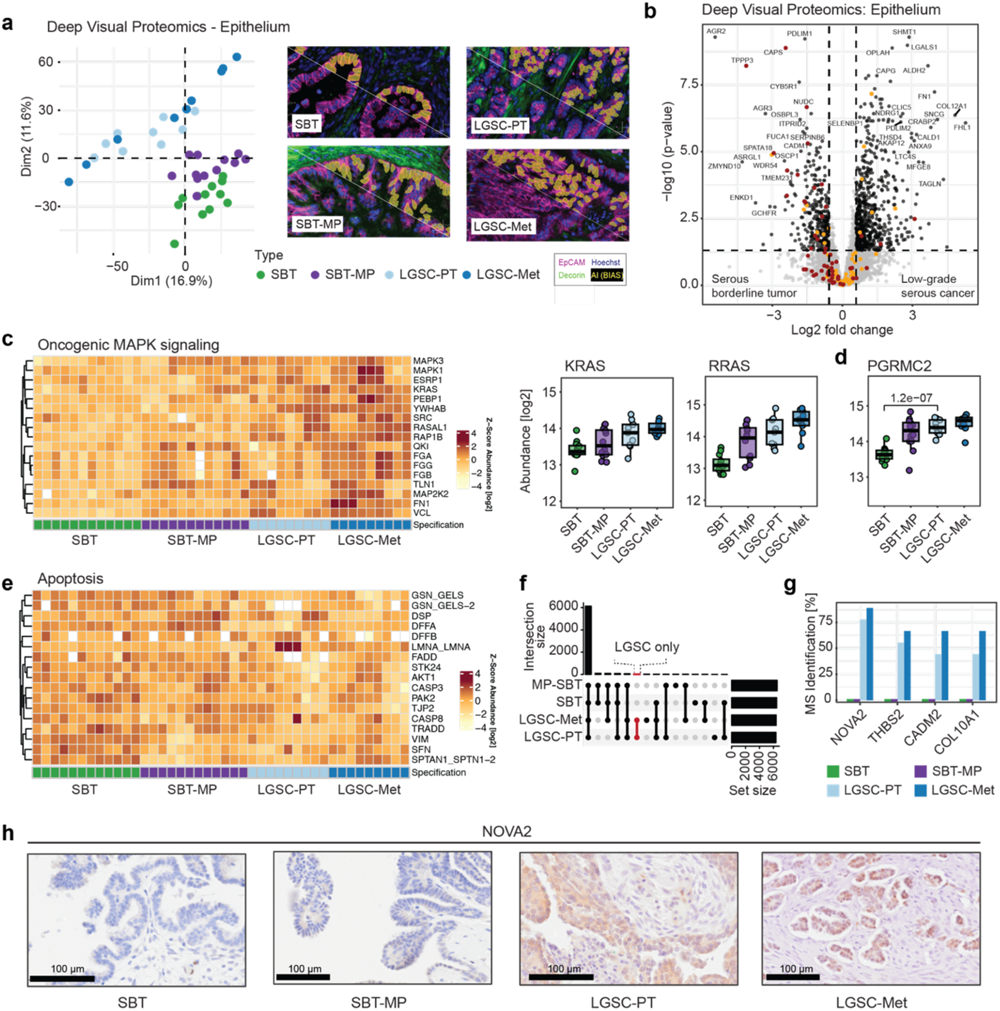
Deep Visual Proteomics on the epithelial tumor compartment confirms known and identifies novel pathways in transition of SBT to LGSC. **a)** Principal Component Analysis (PCA) for the epithelium clearly separates serous borderline tumors, serous borderline tumors with a micropapillary pattern, invasive low grade serous cancer, and corresponding metastasis. This transition is evident in the diagonal of PC1 and PC2 from lower right to upper left. AI-based recognition of epithelial cells using immunofluorescence (EpCAM-purple, decorin-green) below the white diagonal line, followed by AI segmentation (yellow). **b)** Volcano plot of the differential epithelial protein expression between SBT and LGSC-PT in the epithelial compartment. A fold change cutoff of 1.5 and a q-value cutoff of 0.05 are indicated by vertical and horizontal lines, respectively. Proteins matching the significance for differential regulation (DR) criteria are highlighted in black, markers of secretory cells in orange and ciliated cells in red. **c)** Proteins of the mitogen-activated protein kinase (MAPK)-signaling pathway show a gradual increase towards LGSC and corresponding metastasis (Heatmap). Commonly altered Ras and Ras-regulating proteins (box plots). **d)** Boxplots of significantly changed membrane-associated progesterone receptor component 2 (PGRMC2) between the four groups (Student’s T-test). **e)** Heatmap. Proteins involved in apoptosis show reduced abundance from SBT to LGSC-Met. **f)** Upset plot. Comparison of MS-detected peptides/proteins detected in specific groups but completely absent in others and therefore not included in Fig. 1e and 2b (methods). The set size is the number of identified proteins, while the intersection size shows the number of overlapping proteins. **g)** Bar plot of the protein subset highlighted in (f). Bars present the percentage of samples in which the four most frequent proteins were identified per group using mass spectrometry. NOVA2 was solely detectable in more than 75% of LGSC-PT and LGSC-Met, but not in SBT and SBT-MP. **h)** Immunohistochemistry for NOVA2.

Serous borderline tumors with micropapillary features in the background of an SBT or uniform SBT-MP have a higher risk of recurrence as LGSC than conventional SBT^11^. However, whether SBT-MP (Fig. 1a) is a precursor to LGSC is controversial. In our proteomic data, the most significant changes between SBT and SBT-MP included metabolic changes, such as abundance changes in the argininosuccinate synthase (ASS1), a key enzyme in the arginine biosynthetic pathway with ALDH6A1 and ALDH2 (Extended Data 3a). Several proteins showed major expression changes between SBT and SBT-MP, the first step in the hypothesized transition, and maintained their abundance in LGSC and its corresponding metastasis (Extended Data 2a, b, Extended Data 4). We found an increased abundance of the transcription factors AHDC1 and ERF, as well as proteins associated with cancer stemness (AQP5 and ASB6), which were higher expressed in SBT-MP when compared to SBT (Extended Data 4a,b). The tumor suppressor CDKN2A^15^ was already downregulated in SBT-MP and this low protein expression persisted in LGSC and LGSC-Met (Extended Data 4c). This data suggests that SBT-MP protein expression is more similar to SBT than to LGSC, consistent with the PCA (Fig. 2a); however, SBT-MP protein expression reflects several hallmarks of malignant transformation. One patient had both SBT-MP and LGSC adjacent on the same ovarian section, presenting a unique opportunity to study the putative transformation of SBT-MP to LGSC (Extended Data 5a). The gradual progression from SBT to invasive cancer identified in the overall cohort, was confirmed in the PCA of this specific case (Extended Data 5b, c). The differential protein expression mirrored findings obtained by comparing SBT-MP and LGSC in the main cohort, including the downregulation of the ubiquitin ligase, TRIM25, the second most significant protein in LGSC tissue of the main cohort (Extended Data 5d).

A direct comparison of SBT to LGSC-PT in the epithelial compartment identified 963 significantly differentially expressed proteins (Supplementary Table 5). These included several upregulated proteins previously identified in high-grade serous ovarian cancer (SHMT1, TAGLN, Fig. 2b). Two tumor suppressors involved in epigenetic regulation were progressively lost in LGSC and its corresponding metastasis (ZMYND10, OSCP1/NOR1), as was Anterior Gradient Protein 2/3 (AGR2/3) previously implicated in the progression of SBT to LGSC^16^ (Extended Data 2b). Comparing SBT and LGSC-PT, 14.5% of all proteins showed a significantly different abundance, in contrast to only 0.3% between LGSC-PT and LGST-Met, indicating the high degree of similarity between primary tumors and corresponding metastasis at the single cell type level. Neudesin (NENF), involved in the proliferation of neural progenitor cells^17^, was one of the few upregulated proteins in the metastasis (Extended Data 3a). We found a gradual increase in the adipogenesis regulatory factor ADIRF, which is associated with cisplatin resistance^18^, and the folate receptor alpha (FOLR1), which is a target for Mirvetuximab Soravtansine, a treatment of platinum resistant high-grade serous cancers^19^ that is not currently used for LGSC (Extended Data 2a).

The MAPK-signaling pathway has been found to be altered in more than 50% of LGSC and 60% of serous borderline tumors^20^. We found that oncogenic MAPK, ERK1/2 and Met signaling were enriched in LGSC when compared to the borderline tumors (Fig. 2c, Extended Data 3b, c, Supplementary Table 6). Specifically, KRAS and RRAS gradually increased from SBT to LGSC (Fig. 2c), as did downstream effectors of the c-Met receptor tyrosine kinase (Extended Data 3c). Likewise, several proteins downstream of TGF-β signaling were gradually upregulated either in SBT-MP or in LGSC, including oncogenic ADAM17, which promotes epithelial to mesenchymal transition (EMT)^21^ (Extended Data 3d). Most SBT and LGSC express hormone receptors and are at least temporarily responsive to anti-hormonal treatment^22^. Accordingly, there was also an increase from SBT to SBT-MP in progesterone binding protein, PGRMC2, indicating a role for progesterone during the early transition to LGSC (Fig. 2d)^23^.

We also confirmed the previous finding that serous borderline tumors express PAX8, a key marker for secretory cells of the fallopian tube^15^. However, SBT expressed a large number of proteins reflecting normal ciliated cells of the fallopian tube including CAPS, TPPP3 and NUDC^24^, which, interestingly, were lost upon progression to LGSC (Extended Data 3b,e,f, Supplementary Table 6).

Immunofluorescence staining confirmed the proteomics results, detecting only sporadic ciliated cells in LGSC (Extended Data 3f). This was accompanied by a decrease in proteins of the apoptotic machinery, including FADD, CASP-3/8, and TRADD (Fig. 2e). Among proteins identified by MS in LGSC and metastasis but absent in SBT and SBT-MP, we found exclusive expression of the splicing regulator NOVA2, which is physiologically expressed in the brain, but not in the healthy ovary^25^ (Fig 2f, g, Supplementary Table 7). Of note, NOVA2 was detected in the medium abundance range of MS intensities rather than indicating incomplete MS data acquisition by values in the low abundance range, thereby validating our previous proteomic analysis (Extended Data 3g). NOVA2 expression was confirmed using IHC (Fig 2h).

### Spatial proteomic analysis of the tumor microenvironment

Deep Visual Proteomics allows protein detection in different tissue compartments, taking advantage of AI-based recognition of individual cell types (Fig. 3a). When analyzing stromal cells (as opposed to epithelial cells before) which we identified by decorin IF staining, a total of 178 proteins significantly changed during the transition from SBT to LGSC (Extended Data 6, Supplementary Table 4). SBT and SBT-MP global protein expression was clearly distinct from that of LGSC and its corresponding metastasis in a PCA (Fig. 3b), demonstrating that major stromal changes occur as the tumor becomes invasive (Extended Data 7a, Supplementary Table 5). In contrast to the epithelial compartment (Fig. 2a), the stroma of SBT and SBT-MP was very similar, showing no intermediate progression towards LGSC.

**Fig. 3.**
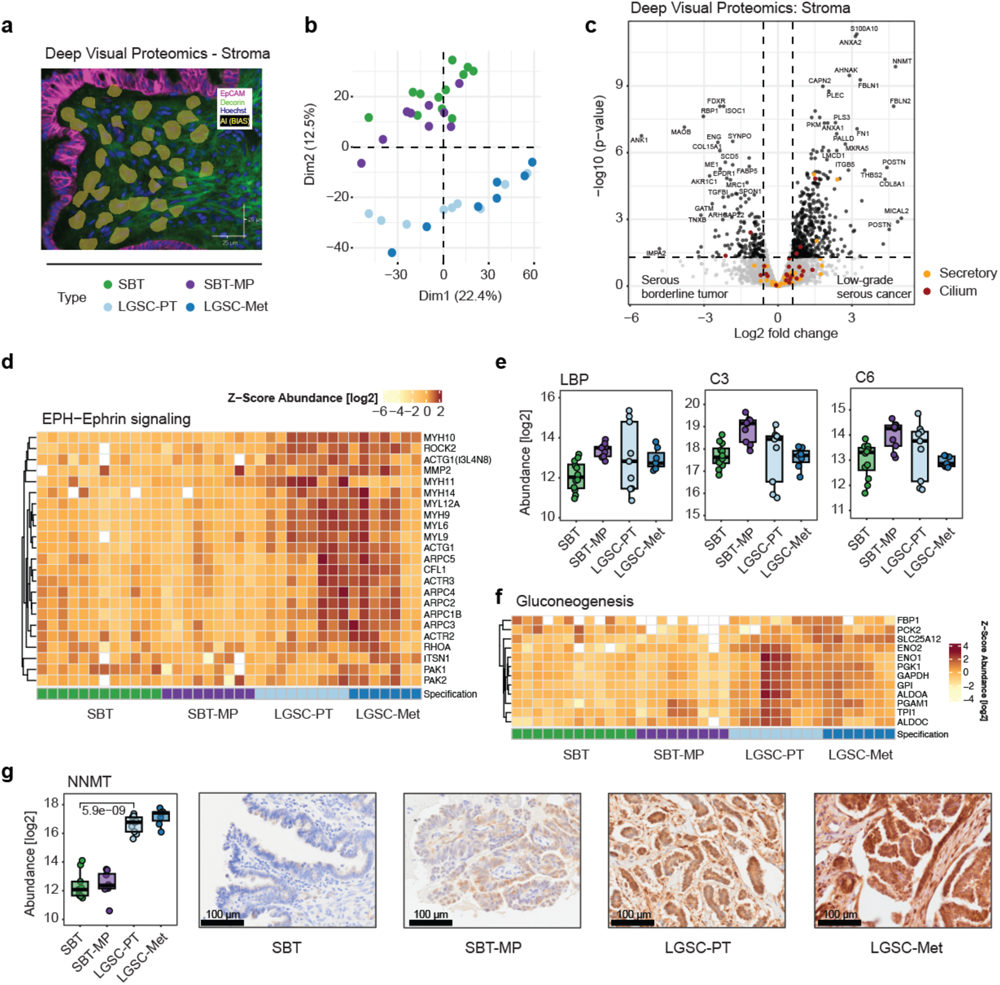
Deep Visual Proteomics of the stromal compartment uncovers a bimodality in the transition of SBT to LGST. **a)** Immunofluorescence outlining the extraction of cell equivalents from the stroma (EpCAM-purple, decorin-green, AI-segmentation - yellow). **b)** Principal Component Analysis comparing stromal protein expression shows the separation of serous borderline and micropapillary tumors from invasive low grade serous cancer and corresponding metastases. The variability was most evident in dimension 2 (12.5%). **c)** Volcano plot of differential stroma protein expression between SBT and LGSC-PT in the epithelial compartment. A fold change cutoff of 1.5 and a q-value cutoff of 0.05 are indicated by vertical and horizontal lines, respectively. Proteins matching the significance for differential regulation (DR) criteria are highlighted in black, markers of secretory in orange and ciliated cells in red. **d)** Proteins that are involved in actin-rearrangement suggested by ephrin (EPH) signaling show an increase towards LGSC and corresponding metastasis in the stromal compartment. **e)** Boxplots of proteins involved in acute inflammation and the complement system across the transition. **f)** Proteins involved in gluconeogenesis show an increase towards LGSC and corresponding metastasis. **g)** NNMT protein abundance (left) and immunohistochemistry (right).

Stromal protein expression changed primarily during the transition from SBT to LGSC (629 differentially expressed proteins), including the upregulation of ANXA2 and its regulator S100A10, which play a central role in cancer cell proliferation (Fig. 3c). Proteins involved in EMT such as AHNAK, LMCD1 and Periostin (POSTN) were among the most significantly differentially expressed stromal proteins of the SBT to LGSC transition. These changes in expression were accompanied by significant alterations in ECM remodeling proteins (PLEC, FBLN1/2, MXRA5, FN1 and its receptor ITGB5), indicating a dense tumor microenvironment.

Biological processes upregulated in LGSC-PT were primarily associated with cell dynamics; we found an upregulation of actin (ACTG1) and its central regulator ROCK2, as well as other components involved in cytoskeletal organization (ARPC2-4, CFL1, ITSN1, PAK1/2), all known to be associated with ephrin signaling (Fig. 3d, Supplementary Table 6). Elastic fiber formation gradually increased towards invasive phenotypes (Extended Data 6b). Similarly, proteins involved in cell-cell adhesion were more abundant in the invasive LGSC and included the suppressors of apoptosis, COMP and GAS6, confirming our findings of decreased apoptosis in the epithelium (Extended Data 7c, Fig. 2e). In addition, we found an acute inflammatory response in micropapillary SBT, which was less evident in the invasive tumors (Fig. 3e, Extended Data 7d). Thus, during early transformation (SBT-MP), our data suggest that there is a strong host immune reaction; however, once an invasive tumor (LGSC) is established, the expression of proteins associated with an immune response is downregulated in the tumor organ.

Our data also suggest that the stromal transition from non-invasive to invasive tumor stages is accompanied by increased glucose metabolism, including the upregulation of FBP1 and PCK2/PEPCK, which are rate-limiting enzymes in gluconeogenesis^26^ (Fig. 3f). The master metabolic fibroblast regulator, NNMT, previously shown by us to regulate the transition from normal fibroblasts to cancer-associated fibroblasts (CAFs)^27^, was among the most highly upregulated proteins in invasive LGSC as confirmed by IHC (Fig. 3g). In the IHC analysis NNMT expression was also high in the epithelium.

### The progression of serous borderline tumor to invasive low-grade serous carcinoma characterized by spatial transcriptomics

Protein expression best reflects the functional phenotype of a cell; however, we reasoned that correlating protein expression to gene expression changes would help us better understand the biology of LGSC. Combining spatial proteomics with spatial transcriptomics could provide information about bidirectional communication between tumor cells and the surrounding stroma and identify treatment targets missed by either method alone^28^.

Using the GeoMx technology, which allowed us to define specific regions of interest (ROIs), we performed spatial transcriptomics of the serial sections of all four histologies that had been used for H&E and spatial proteomics (Fig. 1a, b). Following hybridization with over 18,000 RNA-probes, focused UV light allowed us to selectively release the probe barcodes that hybridized to the cell types of interest for NGS analysis (Supplementary Table 8). Sequencing counts of transcript probes generally indicated high sequencing saturation (9,872 targets detected after QC, Extended Data 8). To integrate the spatial transcriptome with the proteome, we matched ROIs of transcript expression by immunofluorescence staining using compartment specific antibodies (Fig. 4a).

**Fig. 4.**
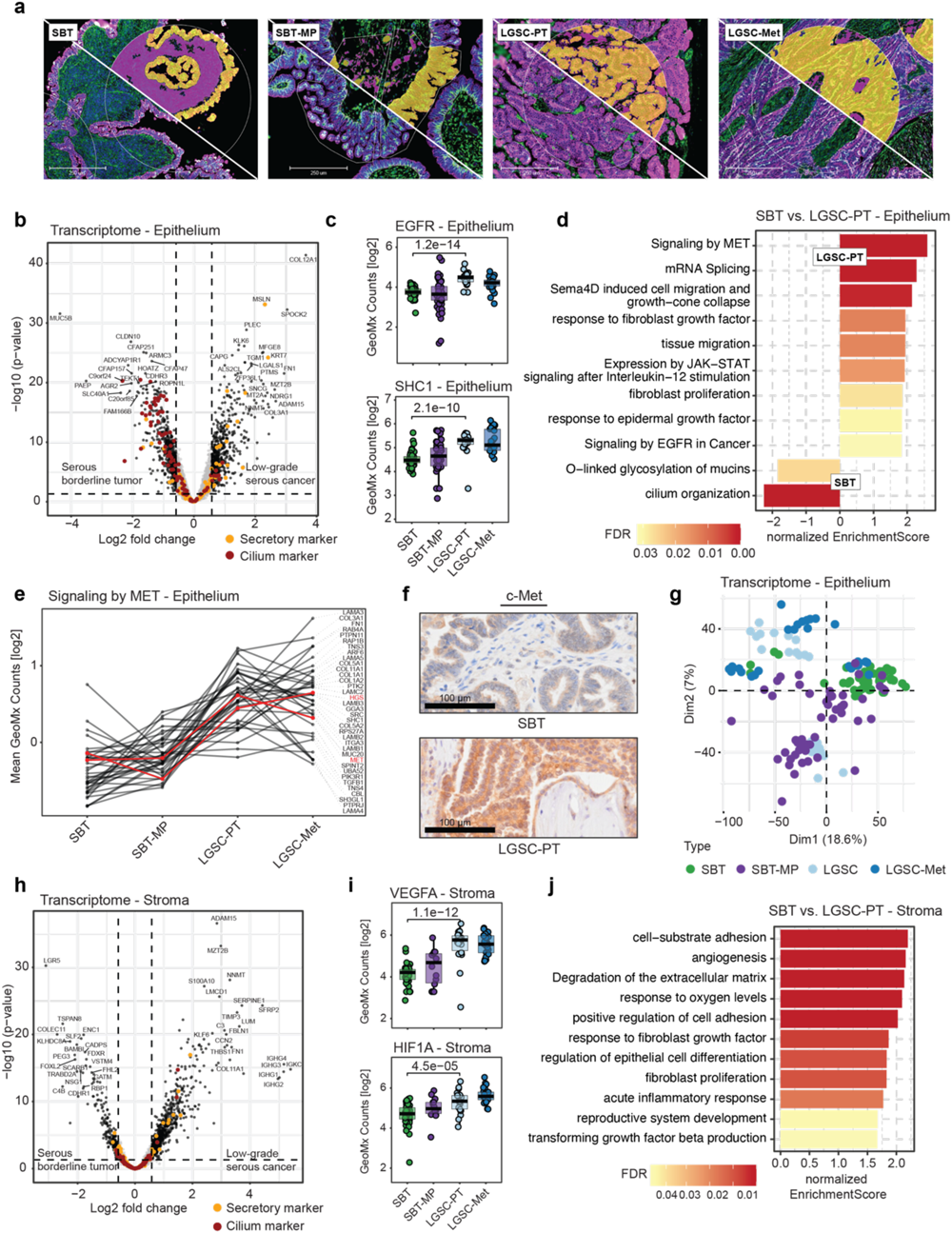
Spatial transcriptomics of SBT and LGSC. **a)** Immunofluorescence for pan-cytokeratin (purple), decorin (green), and nuclei (blue) for exemplary regions. Tumor and stroma compartments for subsequent UV illumination are shown above the white diagonal line in yellow and magenta, respectively. Regions of interest are outlined with fine white lines. **b)** Epithelium. Differential transcript abundance of borderline versus low-grade serous cancer (Volcano plot). Protein markers for ciliated and secretory cells are highlighted in orange and red, respectively. **c)** GeoMx counts for EGFR and SHC1 transcripts across the progression series (Student’s t-test). **d)** GSEA biological pathway enrichment analysis based on the spatial transcript results comparing SBT and LGSC-PT in the epithelial compartment (Pathway REACTOME, Gene Ontology Biological Processes) on the comparison in (b). **e)** Profile plot of pathway-associated proteins determined in (d) for ‘Signaling by MET’. Proteins with critical roles in the pathway (MET, HGS) are annotated in red. All other proteins are summarized in Supplementary Table 8. **f)** IHC of c-MET in SBT and LGSC-PT. **g)** Nanostring Principal Component Analysis using transcripts in the epithelium for the indicated histologies. **h)** Stroma. Differential transcript abundance of SBT versus LGSC-PT. Protein markers for ciliated and secretory cells are highlighted in orange and red, respectively. **i)** GeoMx counts for VEGFA and HIF1α across the progression series (Student’s t-test). **j)** GSEA biological pathway enrichment analysis using the spatial transcript results in SBT and LGSC-PT in the stromal compartment (Pathway REACTOME, Gene Ontology Biological Processes) on the comparison in (h).

A total of 1,386 transcripts in the epithelium were differentially expressed between SBT and LGSC-PT (Supplementary Table 9). In SBT, the mucin-regulating AGR2 and the gel-forming mucin MUC5B were upregulated when compared to LGSC (Fig 4b). More abundant transcripts in LGSC-PT included the CA-125 binding partner mesothelin (MSLN) and the serine protease KLK6, which predicts the recurrence of borderline and low-grade ovarian tumors and is regulated by MAPK^29^. Interestingly, several gene transcripts highly expressed in the nervous system, such as SPOCK2 and gamma-synuclein (SNCG) were upregulated in LGSC-PT. In the proteomics data, gamma synuclein was found to have been upregulated during the early transition of SBT to SBT-MP (Extended Data 2a, 4).

During the step-wise transition from SBT to LGSC, the transcriptional regulator MAZ, which is regulated by MAPK, and the transcription factor DUX4^30^, which is involved in oocyte development, were upregulated early in SBT-MP (Extended Data 9a). The expression of JUNB, FOS, and the Fos-related antigen 2 (FOSL2), in conjunction with their upstream regulators EGFR and SHC1, gradually increased as SBT transitioned through SBT-MP to LGSC (Fig. 4c, Extended Data 9b, c, and Extended Data 10a, b). In contrast, the transcriptional repressor HIC1, which is involved in the stabilization of p53, was downregulated in SBT-MP when compared to SBT (Supplementary Table 9). GSEA pathway analysis highlighted changes in mucins such as MUC3A, MUC5B, and MUC16 (CA-125) as well as tissue migration, semaphorin signaling, and response to EGF (Fig. 4d and Extended Data 9d-f, Supplementary Table 6). Another receptor tyrosine kinase (RTK) receptor, c-Met, and its downstream effectors showed a very significant increase in both LGSC and LGSC-Met (Fig. 4e), which was confirmed by IHC (Fig. 4f) and is consistent with the spatial proteomics results (Extended Data 3c).

PCA analysis of the Nanostring data in the epithelium showed differences between SBT and SBT-MP, while stromal gene expression distinctly separated SBT/SBT-MP from LGSC-PT/LGSC-Met (PCA, Fig. 4g, Extended Data 11b). Several known stromal transcripts upregulated between SBT and LGSC, such as S100A10, C3, and NNMT (Fig. 4h, Supplementary Table 9) had also been identified by spatial proteomics (Fig. 3c, Supplementary Table 5). Others, only identified as transcripts, such as the metalloproteinase, ADAM15, and transcripts involved in oxygen-hemostasis (e.g. VEGFA, HIF1α), increased during the transition from SBT to SBT-MP (Fig. 4i). In the early differences between SBT and SBT-MP, we found an enrichment of the stem cell markers LGR5 and FOXL2 in SBT (Extended Figure 10c,d, Extended Data 11a). The neural axon guidance factor SLIT2 was highly expressed in LGSC-Met, while the cell-adhesion protein CDHR1 was downregulated (Extended Data 11a). Upregulated pathways in CAFs^31^ included genes involved in angiogenesis and hypoxia (Fig. 4j, Extended Data 11c).

### Integration of spatial proteomics and transcriptomics technologies

We reasoned that the multiple spatial data layers – single cell-type proteomics, transcriptomics and H&E staining – might complement each other when trying to understand the tumor organ. In particular, integrating protein and transcript expression could uncover the complex interplay of transcription and translation. Among the proteins and transcripts that did not change between SBT and LGSC-PT (Supplementary Tables 5, 9), 75% of the proteins and 38% of the transcripts overlapped between DVP and Nanostring in the stroma, while 43% of the proteins and 29% of the transcripts overlapped in the epithelium. In contrast, there was less overlap of the differentially expressed proteins and transcripts – 4% of proteins and 2% of transcripts in the stroma as well as 3% of proteins and 2 of transcripts in the epithelium with many Nanostring values not present in the DVP data and *vice versa* (Extended Data 12a, b).

These values reflect differences in the gene products detectable by the two technologies and potential differences in biological regulation^32^. We therefore focused on genes for which both the transcript and the protein were quantified, integrating the two datasets into a list of proteins/transcripts acquired through spatial proteomics and transcriptomics.

To determine markers with a higher probability of impacting tumorigenesis, we selected the most relevant proteins/transcripts acquired with both technologies for epithelium and stroma, respectively (selection scheme in Extended Data 12c). Downregulated targets were included if present in a tumor suppressor database^33^. The resulting list of 70 potential targets was integrated with a comprehensive literature review to annotate for biological relevance (Extended Data 12c-g, Supplementary Table 10). Notably, multiple proteins in both the stroma and the epithelium were targetable with clinically approved or pre-clinical inhibitors. These included CCN2 and ANXA2 in the stromal compartment, FOLR1, CRYAB and HTRA1 in the epithelium, and ADAM15 in both cell types.

Using serial sections for H&E, DVP, and Nanostring allowed perfect alignment of regions of interest from the H&E to transcriptomics and proteomics (Fig. 5a). Correlating the proteome and transcriptome data sets across the four histologies, a total of 1,142 of 4,992 genes present in the epithelium showed a positive correlation across the transition from SBT to LGSC-Met (Fig. 5b). In the stroma, 84 of 3,746 significantly correlated between transcripts and proteins (Fig. 5c). Only very few genes were anti-correlated between the two technologies in the epithelial compartment (total of 32, including ABT1, ZMIZ1, CIR1), and only a single gene in the stroma (CHST14). The majority of the 70 genes of potential biological relevance in the transition from SBT to LGSC selected above showed a similar trend between technologies (Extended Data 12c-g, Supplementary Table 10).

**Fig. 5.**
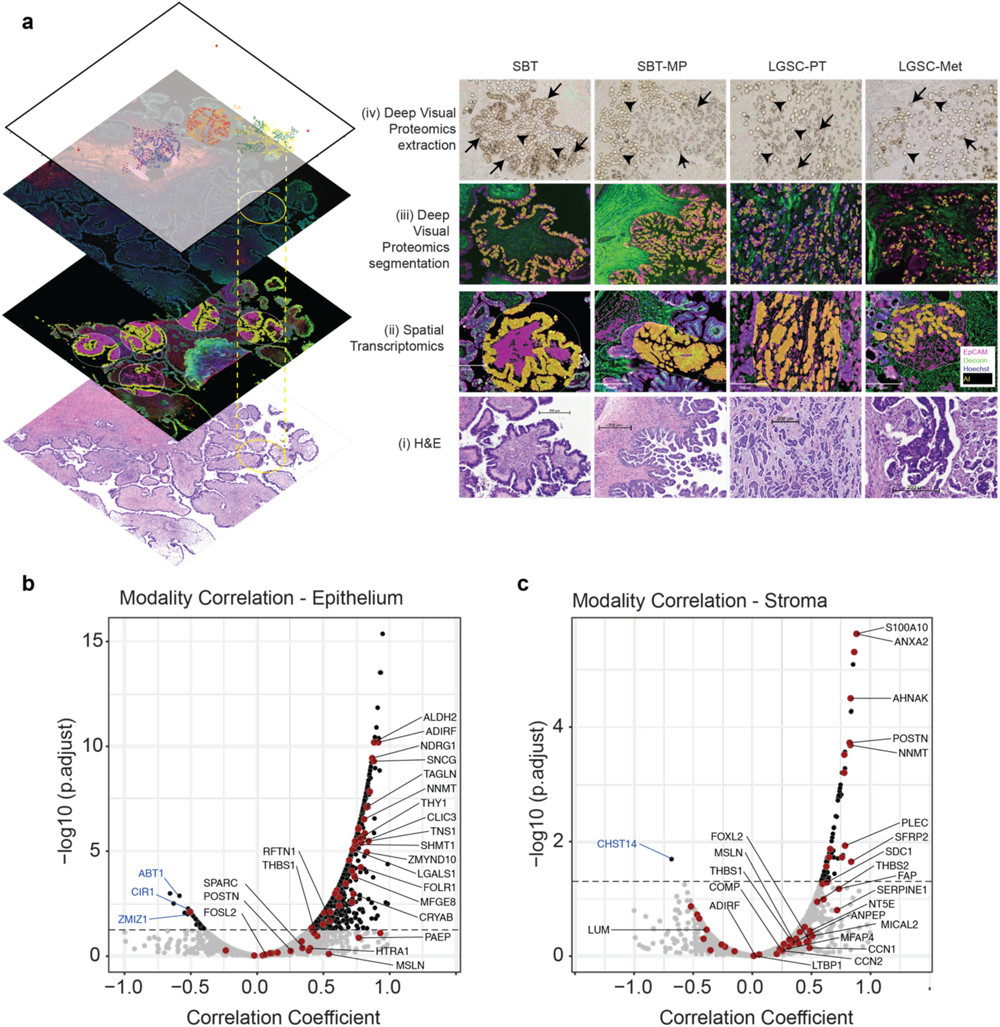
Integration of spatial transcriptomics and proteomics. **a)** Multi-layer integration of Deep Visual Proteomics and spatial transcriptomics for the four histologic subtypes. (i) H&E, (ii) spatial transcriptomics regions of interest, (iii) Deep Visual Proteomics including AI based cell recognition/segmentation, and (iv) brightfield image after DVP laser microdissection. Immunofluorescence showing malignant epithelial cells (EpCAM, purple) and stroma (decorin, green) in both spatial proteomics and transcriptomics (ii, iii). AI-based recognition in the DVP or Nanostring technology is shown in yellow (tumor compartment) and magenta (stromal compartment in spatial transcriptomics), respectively. Microdissected epithelial cells (black arrows) and stroma (arrowheads) (iv). ROIs of spatial transcriptomics matched to the previously selected regions. The top layer of the visual integration (left panel) shows the laser microdissected cells in different regions for the epithelium (blue/yellow/red) and in the stroma (purple/orange/green) used in DVP. **b, c)** Correlation of protein and transcript expression comparing spatial proteomics and transcriptomics. Genes that are significantly correlated are in black and are above the dashed line (Pearson coefficient ≥ 0.05). A negative correlation coefficient indicates opposite trends of protein and transcript expression. Anticorrelating genes discussed in the text are annotated in blue. Significant genes among the set of 70 of biological interest are highlighted in red and annotated in black (see also Extended Data 12).

### From molecular signatures to functional validation: Characterizing the transition of serous borderline tumor to low-grade serous cancer

Having characterized the proteomic and transcriptomic landscape through the progression to metastatic LGSC, we set out to understand the functional significance of the putative drivers of the transition between SBT and LGSC. We used the most significantly altered protein/transcript panel (Extended Data 12c-e, Supplementary Table 10) to evaluate specific alterations from SBT to LGSC. To this end, we first characterized several LGSC cell lines using proteomics and correlated these results with the epithelial compartment acquired with spatial proteomics in human tissue (Fig. 2). All five low-grade cell lines correlated positively with the proteomic findings in the epithelium of SBT and LGSC-PT, with the VOA4627 cell line showing the highest correlation with LGSC-PT (Fig. 6a). Based on these results, a siRNA screen was performed using the VOA4627 LGSC cell line to study migration. Knockdown of ADAM15, CLIC3, NOVA2, POSTN and SNCG, significantly inhibited VOA4627 migration (Fig. 6b). Individual knockdown of all five genes inhibited proliferation of VOA4627 and invasion of VOA4627 and VOA6406 cells, while knockdown of NOVA2 and SNCG significantly inhibited VOA6406 proliferation. (Fig. 6c, Extended Data 13a-c).

**Fig. 6.**
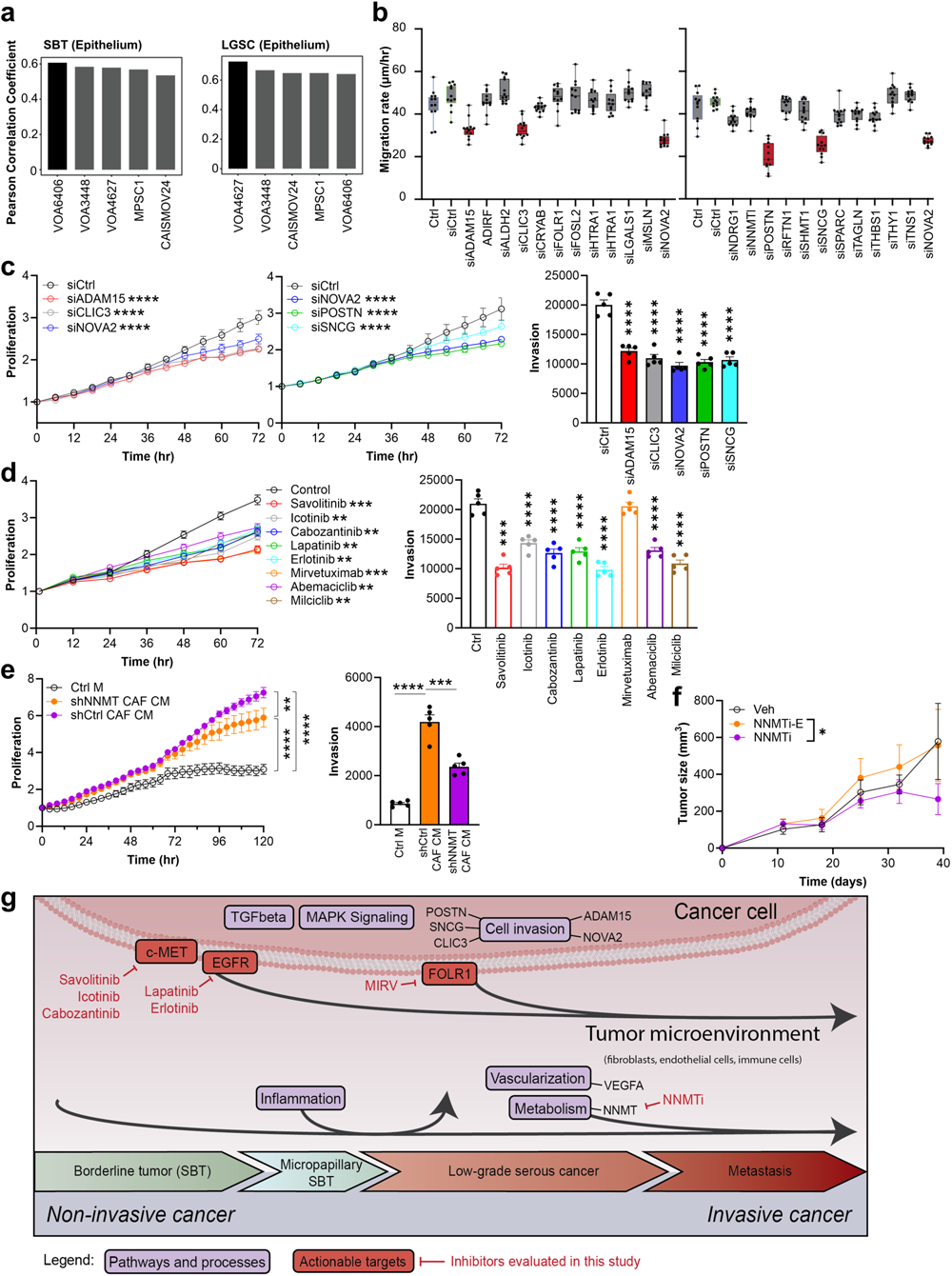
Functional studies characterizing the significant ‘omics’ genes and pathways in the transitions of SBT to LGSC. **a)** Pearson coefficient. Correlation analysis of protein expression (DVP) from SBT (left) and LGSC (right) human epithelial tissue compared to epithelial LGSC cell lines. Cell lines of highest similarity are highlighted in black. **b)** siRNA screening of 23 priority genes (Fig. 5) using the LGSC cell line VOA4627 measuring cell migration. Box plots +/- standard deviation. **c)** Validation of the most promising siRNA hits in the LGSC cell line VOA4627 measuring proliferation (left) and invasion (right). **d)** Inhibitor testing in VOA4627 proliferation (left) and invasion (right). **e)** VOA4627 cells were treated with either condition media from immortalized human CAFs where NNMT was inhibited using shRNA (shNNMT CAF CM), shRNA control transfected CAF condition media (shCtrl CAF CM) or control media (Ctrl M). **f)** Nude mice were injected intraperitoneal with the VOA6406ip1 cell line at day 0. Treatment of mice started at day 11 and was continued daily with either the NNMTi, the inactive enantiomer or vehicle control biweekly and tumor volume measured from 11-40 days after cancer cell injection. All growth curves and bar graphs show mean +/- SEM. The data presented in c-e was repeated in 3 independent experiments. Significance levels by p-value: * 0.05, ** 0.01, ***, 0.001, *** 0.0001. **g)** Schematic summary on pathways and distinct proteins/transcripts involved in the transition from SBT to LGSC.

We explored the role of prominent pathways and targets indicated by our integrative omics approach (Extended Data 12) testing FDA-approved drugs on cancer cell proliferation and the invasion of LGSC cell lines. The c-MET inhibitors, Savolitinib, Icotinib and Cabozantinib, the EGFR inhibitors, Lapatinib and Erlotinib, and the CDK inhibitors, Abemaciclib and Milciclib, all inhibited proliferation and invasion, while a FOLR1-antibody drug conjugate, Mirvetuximab Soravtansine only inhibited the proliferation of VOA4627 and VOA6406 cells, consistent to its mechanisms of action on tubulin which regulates cell proliferation (Fig. 6d, Extended Data 13d). In summary, all pathways projected to be of biological significance from the spatial proteomics or transcriptomics results, turned out to be of functional significance when blocked using FDA approved inhibitors.

NNMT was the most upregulated putative stromal driver in LGSC. Knockdown of NNMT in immortalized human CAF cells^27^ inhibited the CAF-conditioned media-driven proliferation and invasion of VOA4627 and VOA6406 cells (Fig. 6e, Extended Data 13e). A robust and reliable *in vivo* mouse model of LGSC was generated by *in vivo* passaging of VOA6406 cells. Intra-tumoral treatment with an NNMT inhibitor (NNMTi) reduced subcutaneous tumor growth of VOA6406ip1 after 4 weeks of treatment (Fig. 6f).

## Discussion

We performed a detailed analysis of the molecular transition from serous borderline to micropapillary serous borderline tumors and low-grade serous carcinoma and its corresponding metastasis, using cell-type resolved spatial proteomics and transcriptomics. This was enabled by the wider availability of spatial transcriptomics and ongoing improvements of the DVP technology^5^, which now utilizes an enhanced, glycerol-based slide staining protocol^34^, robotic sample preparation, and an increased cutting throughput. Employing this strategy, we identified up to 6,000 proteins in an equivalent of 200 cells in over 100 surgical tissue samples. The tissue-based nature of the DVP and the Nanostring GeoMx spatial transcriptomics platforms permits the use of serial tissue sections for the acquisition of complementary results for the two layers of gene expression and the correlation of the results with H&E staining. Overall, both transcriptomics and proteomics adequately covered previously known biological changes in cancer progression. Currently, transcriptomics provides greater depth, but proteomics was better able to cover changes in the stromal compartment. Proteomics accurately quantified protein level changes during progression, and it is expected that the depth of proteome coverage will increase even further.

Compared to conventional SBTs, serous borderline tumors with micropapillary features (SBT-MP) often involve both ovaries and have a higher risk of recurrence as LGSC^35^. Our proteomic data revealed that SBT-MP is an intermediate stage in the transition between SBT and LGSC, as had been postulated based on histology. In the transition of SBT to SBT-MP, we noted the upregulation of genes involved in tumor progression as well as stem cell markers. At the pathway level, this included MAPK signaling, amino acid metabolism changes, and various transcription factors (AHDC1, MAZ, ERF). While transcript and protein expression in the stromal compartment generally did not change between SBT and SBT-MP, proteomics revealed a sudden increase in proteins associated with an inflammatory response, although this subsided once SBT-MP progressed to an invasive cancer (LGSC, LGSC-Met). We speculate that, during the early phase of transformation, the host immune response is activated but then is abrogated when the tumor becomes invasive. Indeed, DUX4, one of the transcription factors we found upregulated in SBT-MP and maintained in LGSC, promotes cancer immune evasion^36^.

The previous molecular characterization of SBT and LGSC focused mainly on genomic profiling and hormone receptor expression. Those studies consistently highlighted changes in RAS genes, *BRAF*, ERBB2, and NF1, all regulators of the MAPK pathway, which is altered in more than 50% of all LGSC and borderline tumors^20,37^. By extending such studies to the proteomics and transcriptomics levels, our results unraveled changes in other targetable components of the MAPK pathways, from receptor tyrosine kinases to serine/threonine kinase and transcription factors, revealing a gradual activation of the entire MAPK network^38^ as borderline tumors undergo a transformation from SBT to SBT-MP to LGSC, and metastasize. By systematically analyzing the transition from SBT to LGSC-Met and separately analyzing changes in the stroma and epithelium, we expanded these insights into other molecular changes in LGSC beyond MAPK (Fig 6 g). A strength of our analysis was the excision of epithelial tumor cells directly from their microenvironment allowing for cell-type specific resolution. Additionally, our spatial compartment-resolved analysis enabled us to study the stromal compartment in closest proximity to the tumor.

Our data suggest that EMT is an essential mechanism in the transition of SBT to LGSC. Indeed, the transcript and protein signatures in the stroma surrounding LGSC suggest marked ECM remodeling and support the observation that stiffness and a denser microenvironment is often associated with progression^39^. The transition of ductal carcinoma in situ to invasive breast cancer is similarly marked by changes in the EMT and a dense tumor stroma^40^. Beyond recapitulating EMT as tumors dedifferentiate, the spatial omics technologies identified specific, actionable targets in the stroma (e.g. ADAM15, CCN2, ANXA2). The most prominent protein and transcript change was NNMT, which reprograms normal fibroblasts into CAFs^27^. We found that a knock-down of NNMT reduced the proliferation in in vitro experiments and in vivo tumor growth, opening up the possibility of stromal-directed treatment approaches as part of LGSC treatment.

One of our most striking findings in the transition to invasiveness was the appearance of proteins associated with the brain. Specifically, proteomics identified NOVA2, a brain-specific splicing regulator, as highly expressed in LGSC and its corresponding metastasis, but absent in SBT and SBT-MP. Integrating spatial proteomics and transcriptomics, followed by validation and functional studies, identified previously unknown mechanisms for LGSC: CLIC3, POSTN, ADAM15 and SNCG represent targets previously unknown in LGSC. The highly significant proteins identified by both spatial techniques and selected for further studies were further narrowed by an siRNA screen and confirmed by functional assays. Based on our combined in vivo and in vitro results, our data supports further development of c-Met, EGFR, NNMT, and treatment using an FOLR1-specific antibody-drug conjugate in LGSC.

In summary, this first multi-omic spatial atlas of low-grade serous ovarian cancer progression molecularly dissected a previously hypothetical transition pathway, identifying a plethora of functionally important proteins and pathways, and suggested novel treatment targets. By characterizing the molecular landscape of serous borderline tumors and low-grade serous cancer on two omics levels, we provide an in-depth resource for further downstream analyses of the diseases, while demonstrating the translational relevance of our results in*-*vitro and in*-*vivo in this study. The strategy exemplified here could serve as a blueprint for research into other cancers or diseases that would benefit from multi-modal, cellularly, and spatially resolved elucidation of the transition from benign to malignant phenotypes.

## Data Availability

All data will be publicly available upon publication in a journal

## Acknowledgments

We would like to thank Dr. Annette Feuchtinger and her team from the Core Facility Pathology & Tissue Analytics at the Helmholtz Center, Munich, Germany for their support during image data acquisition; Dirk Wischnewski (MPI Munich), Destiny Brown (U of C) and Paul Miller (U of C) for technical support; members of the Department of Proteomics and Signal Transduction at the Max Planck of Biochemistry in Munich, Germany, the ovarian cancer research laboratory in Chicago, USA, as well as members from the Center for Protein Research at Copenhagen University, Denmark, for fruitful discussions. We thank Pieter Farber, Ph.D. from the U of C Genomics facility for help with the spatial transcriptomics. We thank Dr. Mark Carey (The University of British Columbia, Division of Gynecologic Oncology, USA), Dr. Ie-Ming Shih (The Johns Hopkins Hospital, Department of Pathology, USA) and Dr. Fernando Guimarães (University of Campinas, Experimental Pathology Laboratory, Brazil) for low-grade cancer cell lines. We thank Gail Isenberg (U of C) for editing the manuscript.

This research has been supported by generous philanthropic gifts from Linda Usher and family (SD Yamada and E Lengyel), Bears Care - the charitable beneficiary of the Chicago Bears Football Club (H. Kenny and E. Lengyel), and a NIH/NCI R35 CA264619 (E Lengyel). The Cellular Screening Center and Human Tissue Resource Center Cores at the University of Chicago are funded by the Cancer Center Support Grant (P30CA014599). Furthermore, this study was supported by the Max Planck Society for Advancement of Science (M.M.). L.S. acknowledges funding from the German Federal Ministry of Education and Research under the funding code 16LW0243K. L.S. and M.T. were supported by the International Max Planck Research School for Life Sciences – IMPRS-LS. F.A.R. is an EMBO postdoctoral fellow (ALTF 399-2021) and T.M.N. is supported by a Swiss National Science Foundation (SNSF) Early Postdoc Mobility (P2ZHP3-199648) and Postdoc Mobility Fellowship (P500PM_210917). M.T. was supported by European Union’s Horizon 2020 research and innovation program under grant agreement No. 874839 (ISLET).

## Contributions

The study was conceived by E. L. and M.M. Deep Visual Proteomics (DVP) application, data acquisition and interpretation were performed by L.S., R.K. and A.M. with the support of M.T., T.M.N., F.A.R., E.R. and A.B. Spatial transcriptomics data were acquired by R.K., L.S., A.B. and S.N.B. Omics data were statistically evaluated and integrated by L.S., M.S., M.C., and J.B.M-R. The patient cohort was identified by E.L., R.M., S.D.Y, R.K, and R.R.L. Pathological expertise was provided by R.M., R.R.L. and A.B. Functional experiments were designed by A.S., H.K., L.S., J.H., and E.L., and performed by A.S., H.K., S.R., L.K. and J.H. Figures were prepared by L.S. and H.K. The paper was written by L.S. and E.L. The manuscript was edited by H.K., A.M., and M.M. All authors reviewed, provided feedback, and approved the final version of the manuscript.

## Data availability

All data will be publicly available upon publication in a journal.

## Methods

### Patient cohort and ethics

Patients who underwent primary surgery for a newly diagnosed borderline ovarian cancer or low-grade serous ovarian cancer at the University of Chicago were retrieved from the University of Chicago ovarian cancer database^42^. All patients gave written informed consent in compliance with the University of Chicago Institutional Review Board-approved protocol and in accordance with the Declaration of Helsinki. All surgeries were performed by board certified gynecologic oncologists (S.D.Y, E.L). The tumor pathology was reviewed and confirmed by two gynecologic pathologists (R.R.L., R.M.) prior to inclusion in the study.

### Deep Visual Proteomics

#### Immunofluorescence

Formalin-fixed paraffin embedded (FFPE) tissue was sectioned (2.5 μm) from paraffin blocks and mounted on 1.0 PEN membrane slides (MicroDissect, MDG3P40AK). To enhance tissue adhesion, membrane slides were incubated for 1 hour under UV light and treated with VECTABOND reagent (Biozol, VEC-SP-1800) according to the manufacturer’s instructions. Mounted slides were incubated at 55°C for 30 min and deparaffinized through xylene (2 min) and 100% ethanol, 95% ethanol, 75% ethanol, 30% ethanol, and distilled water (1 min) two times. Antigen retrieval occurred in 1x DAKO pH9 HIER buffer (Agilent Dako)/ 10% glycerol (v/v) (Sigma) in a preheated water bath at 90°C for 60 min, followed by the blocking of nonspecific binding sites using 5% BSA in PBS for 60 min at room temperature.

Directly conjugated antibodies targeting EPCAM (Abcam, recombinant Alexa Fluor® 555, ab275122, 1:100) and Decorin (Abcam, recombinant Alexa Fluor® 647, ab28132, 1:100) were diluted in antibody diluent solution (Agilent Dako, S080983-2) and incubated at 4°C overnight in a wet staining chamber. Nuclei were stained with SYTOX^TM^ green (1:400, Thermo Fisher Scientific) for 10 min at room temperature and mounted with aqueous mounting medium (SlowFade™ Diamond Antifade Mountant, Thermo Fisher Scientific).

#### Image acquisition and analysis

High-resolution fluorescence images were acquired on an Axio Scan.Z1 (Zeiss) microscope, coupled to a 20x/0.8 M27 dry objective and the scanned slides saved as ’.czi’ files. Images were recorded implementing a 10% tile overlap, five z-stacks (offset -5um) and a bin mode of 1x.1, using optimized exposure times for each fluorescent channel (AF647: 40ms, AF555: 300ms, AF488: 10ms). For image post-processing, z-planes were collapsed into a single plane based on the variance of pixel values (‘Extended Depth of Focus - EDF’) and stitched to achieve precise matching of tiles. Then, images were imported into the Biology Image Analysis Software (‘BIAS’) and analyzed^14^. Briefly, segmentation was performed for epithelial cells (Algorithm: ‘Generic cytoplasm segmentation v1.0’, Settings: input spatial scaling: 1.0, detection confidence: 50%, contour confidence: 50%, region properties: 10-500 μm^2^) and stromal cell equivalents (Algorithm: Generic nucleus segmentation v1.0. Settings: input spatial scaling: 2.4, detection confidence: 50%, contour confidence: 50%, region properties: max. 100 μm^2^, dilated by 9 μm^2^).

Once regions of interest were selected using matching regions in spatial transcriptomics, shapes of single cells were exported while defining three reference points for coordinate system transfer. To improve the efficiency of laser-guided shape extraction, polygon reduction was accomplished by implementing the Ramer-Douglas-Peucker algorithm. To facilitate this, an interactive web interface was developed using Python (version 3.8.5) in conjunction with the Streamlit library (version 1.19.0). Internally, data manipulation tasks were carried out using the numpy (version 1.22.2) and pandas (version 1.4.0) libraries. Visualization of both original and reduced shapes was performed using the plotly library (version 5.5.0). Epsilon values were chosen interactively to find an optimum to preserve shapes and reduce points. Upon image acquisition, cover glasses were removed and the tissue dried thoroughly at room temperature to enable precise laser cutting thereafter.

#### Laser microdissection

Cells were cut from the tissue using the laser microscope (Leica Microsystems) and collected into a dry 384-well plate (Eppendorf) while maintaining a stable temperature of 31.9°C. AI-defined shapes of cells were imported using the reference points defined in the BIAS software and minimal correction of shape alignment was performed. Laser extraction was performed directing a diode-pumped solid-state laser (349 nm) *via* a HC PL FLUOTAR L 63x/0.70 objective (power: 59, aperture: 1, speed: 20-25, head current: 42-49%, pulse frequency: 2450-2600, offset: 214-219) conducting a final middle pulse to collect the shapes vertically into the well. Considering the surface area of the cell as indicator of final protein amounts injected into the mass spectrometer, a total of 700 epithelial cell shapes and 150 stroma equivalents were collected to compensate for differences in area sizes.

#### Sample preparation

All laser dissected samples in the 384-well plate were processed in parallel by the implementation of an automated liquid handling platform (Agilent Bravo). Extracted cells were concentrated at the bottom of each well by addition of 28 μL of 100% acetonitrile, centrifugation at 2,000 g for 10 min and vacuum evaporation for 15 min (60°C). Cells were lysed in 4 μl of 60 mM triethylammonium bicarbonate (TEAB) in H_2_O at 95°C for 60 min. After the addition of 1μL of 60% acetonitrile (final concentration of 12% (v/v)), the samples were incubated at 75°C for 60 min. Proteins were digested sequentially, adding 1 μL of 4ng/μL LysC (Wako, 129-02541; in 60 mM TEAB, 12% I) for 3 hours and 1.5 μL of 4 ng/μL trypsin (Sigma-Aldrich, T6567; in 60 mM TEAB, 12% I) overnight at 37°C. The enzymatic digest was quenched using a final concentration of 1% (v/v) trifluoroacetic acid (TFA), centrifuged for 5 min at 1,000g and vacuum dried at 60°C. Samples were resuspended in 2% acetonitrile (v/v), 0.1% trifluoroacetic acid (v/v) and the entire volume injected for MS data acquisition.

#### Mass spectrometry measurements and data processing

The LC system of choice was an EASY nanoLC 1200 (Thermo Fisher Scientific). Peptides were separated on a 50 cm in-house packed HPLC column^43^ (75 um inner diameter packed with 1.9 um ReproSil-Pur C18-AQ silica beads (Dr. Maisch GmbH)) with a linear gradient of 120 min from 3 to 30% buffer B in 95 min, followed by an increase to 60% for 5 min, washed at 95% buffer B for 10 min and re-equilibration for 10 min at 5% buffer B (buffer A: 0.1% formic acid (FA) and 99.9% ddH2O; buffer B: 0.1% FA, 80% ACN, and 19.9% ddH2O). The flow rate was kept constant at 300 nl/min, and the column was heated to 60°C by an in-house manufactured oven. The EASY LC system was coupled to a timsTOF SCP mass spectrometer (Bruker) via a nanoelectrospray ion source (Captive spray source, Bruker). The mass spectrometer was operated in dia-PASEF mode using the 16 diaPASEF scan acquisition scheme (standard scheme)^44^. The method covered a m/z range from 400 to 1200 and ion mobility of 0.6 to 1.6 Vs cm^-2^. All other settings were described previously^45^. For cell line experiments, the LC system was coupled to a timsTOF Pro2 mass spectrometer (Bruker) with settings as described above.

Raw data was searched using the DIA-NN software^46,47^ (version 1.8.0). Searches were performed separately for the epithelial and stromal compartments using the library-free search and the human Uniprot databases (UP000005640_9606 with isoforms, February 28, 2022). In short, a deep-learning module, match-between-runs (MBR) and heuristic protein inference (‘--relaxed-prot-inf’) was enabled. N-terminal methionine excision and carbamidomethylation were set as fixed modifications, ‘IDs, RT & IM profiling’ was used for library generation, ‘robust LC (high accuracy)’ for quantification and ‘Global’ for cross-run normalization. The ‘pg_matrix.tsv’ output file was used for further data analysis.

### Bulk proteomics of cell lines

The proteome of five cell lines was measured. Cells were harvested and washed in PBS. Pellets were lysed for 10 min at 90°C in 50 uL lysis buffer (12.5% acetonitrile, 300 mM Tris/HCl pH 8.0, 5 mM TCEP, 25 mM CAA), sonicated in a Bioruptor Plus (Diagenode, sonication cycles of 30 sec for 15 min) and heated at 90°C for 30 min. Protein concentration was measured using a Nanodrop instrument (ThermoFisher Scientific), proteins were digested using LysC and trypsin in a ratio of 1:100 over night at 37°C. Digests were quenched by adding trifluoroacetic acid (TFA) to a final concentration of 1% and peptides purified using two layers of SDB-RPS as active matrix in stage tips (Wash 1: isopropanol, 1% TFA; Wash 2: 0.2% TFA; elution buffer (80% I, 1% NH_4_+)). Purified peptides were vacuum dried for 40 min at 60°C and resuspended in 2% acetonitrile (v/v), 0.1% trifluoroacetic acid (v/v). A total of 200ng were injected into the mass spectrometer and measured as described above.

### GeoMx Spatial Transcriptomics

FFPE tissue sections (5 μm) sliced consecutively to Deep Viusal Proteomics samples were processed following the Nanostring GeoMx user manual (MAN-10132-04). In brief, tissue slides were deparaffinized and rehydrated, followed by protein target retrieval using antigen retrieval buffer (Tris-EDTA) for 20 min in a pressure cooker. RNA target retrieval was accomplished by a digest with proteinase K for 15 min at 37°C. Tissue was post-fixed in 10% NBF, followed by an overnight hybridization at 37°C with the RNA probes (Human Whole Transcriptome Atlas, 18,000 protein-coding genes coupled to UV-cleavable oligonucleotide barcodes). Excess probes were removed the next day by washing the samples twice in stringent washes at 37°C, followed by an incubation period in blocking buffer W (Nanostring LOT#:2-23020032). Thereafter, tissue was stained using conjugated primary antibodies targeting pan-cytokeratin (mouse monoclonal antibody, Novus NBP2-33200AF488) and vimentin (mouse monoclonal IgG1κ, Santa Cruz sc-373717) as well as the SYTO13 nuclear stain (ThermoFisher Scientific, S7575) for one hour at room temperature. For transcriptomics data acquisition, slides were placed on the GeoMx Digital Spatial Profiler (DSP) and scanned in 20x magnification. Based on immunofluorescence images, regions of interest (ROI) were collected under supervision of an experienced pathologist (AB) and matched to the regions selected in DVP. Upon ROI selection, oligonucleotide barcodes were collected into a 96-well plate by UV-ablation while precisely separating compartments in each region. Sequencing libraries were prepared using the Illumina TruSeq technology for the ligation of sequencing adaptors and amplification. Amplified libraries were purified using two rounds of Ampure XP magnetic bead cleanup (ratio 1.2:1 (beads: library)) (HighPrep PCR from MAGBIO (Cat#: AC-60500). Purified libraries were sequenced in an Illumina NovaSeq flowcell PE50 at a depth of 100x /μm2.

### Data analysis

Bioinformatics analyses of Deep Visual Proteomics and spatial transcriptomics data was performed in the R statistical environment (version 4.2.2). Protein intensities and GeoMx counts were log2 transformed and evaluated alongside as described below. For each dataset, samples belonging to the main cohort were selected, while addition samples (‘case studies’) were processed separately. For proteomic quality control, data completeness was determined by calculating the number of regions (0-3) in which a protein was consistently identified across a specimen. Coefficients of variations (CVs) were determined on non-logarithmic data and complete MS intensities (no ‘NA’ values) (i) within a specimen (intra-specimen) by calculating the variation between different regions and the mean CV values for each group and (ii) between specimen (inter-specimen) by calculating mean MS intensities of regions for each specimen and the CVs of mean region values within each cohort group.

GeoMx raw count data was processed and normalized using the R GeoMxTools package [1]. Segments with fewer than 1000 raw reads, below ∼75% for % Aligned, ∼80% for %Trimmed and Stitched sequencing reads, were removed. We also removed segments with sequencing saturation <50%, negative count < 1 and No Template Control count > 9000. A probe is removed globally if the geometric mean of that probe’s counts from all segments divided by the geometric mean of all probe counts representing the target from all segments is less than 0.1 or the probe is an outlier according to the Grubb’s test in at least 20% of the segments. Segments with less than 5% of the genes detected were removed and genes detected in at least 5% of the segments were kept. Counts were then normalized by Q3 normalization.

Differential protein/transcript abundances of both datasets were determined using the ‘Limma’ package (v3.54.2) using the false discovery rate (FDR) for multiple testing correction and fixed parameters (adjusted p-value <= 0.05 and logarithmic foldchange > 1.5) for the assessment of significance. Likewise, ‘Limma’ was used for regression analyses across groups using a linear model fit followed by empirical Bayes statistics, while the same significance cutoff was used as described above. An overlap of MS protein identification was determined counting ‘NA’ and valid values per group and a combination matrix for upset plots, as well as all heatmaps, were generated using the ComplexHeatmap package (v2.14.0). For all Principal Component Analyses (PCA) and omics data integration, data were filtered per protein for valid values in 2 out of 3 regions in all specimens of at least one group. Imputed sample-from normal distribution (*width of 0.3, downshift of 1.8*). PCA were performed using the FactoMineR package (v2.8). Biological pathway enrichments were accomplished by a gene set enrichment analysis (GSEA) on significantly differentially regulated proteins/transcripts using the WebGestalt gene set analysis toolkit (v0.4.5) in reference to the ‘Reactome Pathway’ and ‘non-redundant geneontology Biological Process’ databases and a Benjamini-Hochberg FDR correction (cutoff: 0.05). Ciliated and secretory cell markers were extracted from the top 100 abundant transcripts of the ciliated and secretory cells in the post-menopausal fallopian tube as presented in recently published work^24^. Remaining plots were generated using the ggplot2 package (v3.4.2) and mean comparisons of p-values were added using paired Student’s t-tests and equal variances. For the integration of omics data, regions of interest were filtered for dataset counterparts and protein/transcript names were matched by first prioritizing non-isoform proteins and then prioritizing the percentage of valid (not ‘NA’) values when selecting between isoforms. If two ROIs of the spatial transcriptomics matched one DVP region, mean GeoMx counts were calculated. Previously imputed data were used for the proteomic dataset. Correlation between datasets was tested by Pearson correlation using the ‘stats’ package (v4.2.2) and Benjamini-Hochberg (FDR) correction. Stacked image creation of our multi-omics approach was done with a custom Python script (version 3.10.11) and Adobe Illustrator (27.5) using the following libraries: pandas (1.5.3), untangle (1.2.1), numpy (1.24.3), PIL (9.4.0) and matplotlib (3.7.1). Initially, the shape data was superimposed on raw slide images, followed by manual adjustments as necessary. Subsequently, each color layer was exported separately as an overlay and integrated into Adobe Illustrator. Schematics were created with BioRender.com (Figure 1) and Adobe Illustrator (Figure 6).

### Antibody-based validation staining

#### Immunohistochemistry

FFPE tissue was sectioned (5 μm) on SuperfrostTM Plus Microscope Slides (Fisher Scientific, 22-037-246). Then, the immunohistochemistry was performed on Leica Bond RX automated stainer. After the standard procedures for deparaffinization and rehydration, tissue sections were treated with Proteinase K (Agilent, S3004) for 5 minutes pre-treatment at room temperature. Anti-NOVA2 [(Novus Biologicals, # NBP1-92196, 1:150], c-MET [Invitrogen, #18-7366, 1:150], NNMT [Santa Cruz Biotechnology, #sc-376048, 1:100], PGRMC2 ThermoScientific, #PA5-59465,1:200], FOLR1 [ThermoScientific, #PA5-24186, 1:200], JUN [Cell Signaling technology, #9165, 1:200], EGFR [ProScience, #33-350, 1:25] was applied on tissue sections for 60 minutes incubation at room temperature. The antigen-antibody binding was detected by Bond Polymer Refine Detection (DS9800, Leica Microsystem). Tissue sections were briefly immersed in hematoxylin for counterstaining and were covered with cover glasses. These slides were imaged using Olympus VS200 Slideview. Images for publication were exported from QuPath version 0.3.2.

#### Immunofluorescence

FFPE tissue was sectioned (5 μm) on SuperfrostTM Plus Microscope Slides (Fisher Scientific, 22-037-246) and deparaffinized as described above (see section ‘Deep Visual Proteomics’). For antigen-retrieval, samples were heated in a water bath at 90°C for 30 min in 1x DAKO pH9 HIER buffer (Agilent Dako) and blocked using 5% BSA in PBS for 45 min at room temperature. Primary antibodies for PAX8 (Mouse mAb 28556, Cell Signaling Technology, 1:400) and CAPS (Rabbit polyclonal PA5-60401, Thermo Fisher Scientific, 1:50) were diluted in antibody diluent solution (Agilent Dako, S080983-2) and incubated at 4°C overnight in a wet staining chamber. Secondary antibodies (anti-Mouse IgG1 AF647, A-21240, Thermo Fisher Scientific, 1:200; anti-rabbit IgG AF750, ab175735, Abcam, 1:200) and ConcavalinA conjugated to tetramethylrhodamine (C860, Thermo Fisher Scientific, 1:150) were incubated for 60 min at room temperature. Nuclear staining was performed using SYTOX^TM^ green (1:400, Thermo Fisher Scientific) for 10 min at room temperature and slides were mounted (Vectashield Vibrance Antifade Mounting Medium, Vector Laboratories, H-1700). The resulting staining was visualized on a Axio Scan.Z1 (Zeiss) microscope as described above (see section ‘Deep Visual Proteomics’).

### Cell lines and culture conditions

VOA3448, VOA4627, and VOA6406 (M. Carey, University of British Columbia) cells were cultured in a 1:1 mixture of MCDB 105 (Cell Application Inc.) and Medium 199 (Gibco), supplemented with sodium bicarbonate (Corning), L-glutamine (Corning), and 10% fetal bovine serum (FBS). The cells were maintained at 37°C in a humidified incubator at 5% CO2. The cell lines were banked in liquid nitrogen and one vial of each passage were confirmed *Mycoplasma* negative using the STAT-Myco kit. The cell lines were validated using short tandem repeat DNA fingerprinting with the AmpFℓSTR Identifier kit and compared with known fingerprints by IDEXX BioAnalytics Laboratories (Columbia, MO). Cells were passaged 2–10 times after thawing before commencing with experiments.

### siRNA transfection

For transient transfections, LGSC were transfected with 25 nM small interfering siRNA pools using DharmaFECT 1 (Horizon Discovery) in M105:M199 media without FBS. Each pool contained four siRNA sequences for *ADAM15* (L-004505-00), *ADIRF* (L-012306-01), *ALDH2* (L-009766-00), *CLIC3* (L-011805-00), *CRYAB* (L-009743-00), *FOLR1* (L-010403-00), *FOSL2* (L-004110-00), *HTRA1* (L-006009-00), *LGALS1* (L-011718-00), *MFGE8* (L-021466-00), *MSLN* (L-006346-00), *NDRG1* (L-010563-00), *NNMT* (L-010351-00), *NOVA2* (L-012590-00), *POSTN* (L-020118-00), *RFTN1* (L-023452-02), *SHMT1* (L-004617-00), *SNCG* (L-011396-00), *SPCARC* (L-003710-00), *TAGLN* (L-003714-00), *THBS1* (L-015337-00), *TNS1* (L-009976-00), ON-TARGET plus Non-targeting siRNA #1 (D-001810-01), or siGLO RISC-Free Control regent (Horizon Discovery; Catalog number; D-001600-01-05).

### Inhibitor testing

The MET inhibitors, Savolitinib (HY-15959), Icotinib (HY-15164A) and Cabozantinib (YT-13016) and the EGFR inhibitors, Lapatinib (HY-50898) and Erlotinib (HY-50896) were purchased from MedChem Express. The FOLR1-targeted antibody drug target, Mirvetuximab was purchased from ImmunoGen, Inc. The CDK inhibitors, Abemaciclib (LY2835219) and Milciclib (PHA-848125) were purchased from SelleckChem. The inhibitors were tested in proliferation and invasion assays using IC50 doses.

### Quantitative PCR with reverse transcription (RT-qPCR)

RT-qPCR was conducted by StepOnePlus Real-Time PCR System (Applied Biosystems). Total RNA was extracted using TRIzol (Invitrogen) and transcribed into cDNA using TaqMan RNA Reverse Transcription Kit (Applied Biosystems). A TaqMan endogenous control was used to normalize mRNA expression. Each PCR assay was performed in triplicate, and relative levels of *NOVA2* expression were calculated using the 2^−ΔΔ^*C*_t_ method.

### Functional assays

#### Migration/siRNA screen

Ovarian cancer cells were plated in 96-well plates (17,000 cells/well) and reverse transfected with target or control siRNA. The wells were washed thoroughly with phosphate buffered saline (PBS) to remove detached cells and growth media added to each well before scratching with a wound making machine. The cells were monitored on the IncuCyte® Live Cell Analysis System (Sartorius), with the wells imaged every hour to evaluate migration.

#### Proliferation

Ovarian cancer cells were plated onto 96-well plates (10,000 cells/well) and incubated for 24 hours to allow cells to adhere before media changed. For siRNA testing, the cells were pre-transfected with target or control siRNA 24 hours prior to assay and treated with growth media. For inhibitor testing, the cells were treated with IC50 concentration of inhibitors in growth media. For conditioned media testing, 24h conditioned media was collected from primary human CAFs that stably expressednon-targeting shCtrl (5′-GCAGTTATCTGGAAGATCAGG-3′) or shNNMT (5′-GCTACACAATCGAATGGTT-3′) as previously described27. The LGSC cells were treated, incubated at 37°C with 5% CO2, and monitored on the IncuCyte® Live Cell Analysis System (Sartorius), with the wells imaged every 4-12 hours to evaluate proliferation.

#### Invasion

Ovarian cancer cells were plated in the top well of the QCMTM 96-Well Cell Invasion Assay plate (20,000 cells/well). For siRNA testing, the cells were pre-transfected with target or control siRNA 24 hours prior to assay and plated in serum-free media. For inhibitor testing, the cells were treated with IC50 concentration of inhibitors in serum-free media. Growth media was placed in the bottom chamber for the siRNA and inhibitor testing. For conditioned media testing, conditioned media was collected from primary human CAFs that stably expressed shCtrl or shNNMT as described above. The LGSC cells were plated in CAF growth media (control media), and in the bottom chamber CAF control media, shCtrl-expressing CAF conditioned media, or shNNMT-expressing CAF conditioned media, was added. The cells were incubated for 48 hours to allow the cells to invade through the chamber. The invaded cells were detached, lysed, and stained with CyQuant GR Dye according to manufacturer instructions. Total fluorescence (480/520 nm) was acquired on a fluorescent plate reader (SpectraMax iD5).

### Western blot analyses

LGSC cells were plated at a concentration of 4 × 10^5^ cells per well to a 6-well plate and cultured for 24-48 hours. The cells were then lysed with an SDS lysis buffer containing 4% SDS and 10 mM HEPES, at a pH of 8.5. Next, 15-25 μg of each cell lysate was separated by SDS-PAGE using 5–20% gel (BioRad) and transferred to a nitrocellulose membrane (Thermo Fisher Scientific). The membrane was blocked with 5% non-fat dry milk (NFDM) (Lab Scientific) in Tris-buffered saline with Tween 20 (TBST) for 1 hour at room temperature The membranes were incubated with primary antibodies against NOVA2 (1:1000) and GAPDH (1:1000) (Proteintech) in 2% bovine serum albumin (Sigma-Aldrich) in TBST at 4 °C overnight. After washing with TBST, the membrane was incubated with secondary antibodies (Cell Signaling) conjugated to horseradish peroxidase at 1:2000 dilution in 5 % NFDM/TBST for 1 hour at room temperature. The proteins were visualized with Clarity Western ECL Substrate (Bio-Rad) or SuperSignal West Femto Substrate (Thermo Fisher Scientific) under ChemiDoc XRS+ System (BIO-RAD). Full-length immunoblots are available in Supplementary Data.

### In vivo xenograft model

*Development of LGSC xenograft model*. Female HSD:Athymic Nude-Foxn1^nu^ (athymic nude; #069(nu)/070(nu/+) mice at age 5-6 weeks and approximately 20 grams were purchased from Charles River. All procedures involving animal care were approved by the Committee on Animal Care at the University of Chicago. Mice were irradiated and injected intra-peritoneally (i.p.) with 5 million VOA6406 cells. The mice were sacrificed 12 weeks post cancer cell injection. The omental tumors were collected, minced into 1mm^3^ pieces, and digested with 2.5U/ml dispase II (17105041, Gibco) at 37 °C and 5% CO2 for 30 minutes. Single cells were collected by 70 µm filtration, washed twice in PBS, and plated in growth media. After a 30-minute incubation, the unadhered cells were collected and plated in a new flask. These mouse tumor derived cells, VOA6406ip1, were passaged 10 times and validated using short tandem repeat DNA fingerprinting with the AmpFℓSTR Identifier kit and compared with known fingerprints by IDEXX BioAnalytics Laboratories (Columbia, MO).

*NNMTi intervention study experiment.* Five million VOA6406ip1 cells were subcutaneously injected with 50 µl of Matrigel (354262 Corning) into 6-week-old athymic nude mice. The mice that developed subcutaneous tumors were randomized. For treatments, 100 µg NNMTi, 100 µg of the less active enantiomer of the NNMTi, or no drug (vehicle control) in 50 µl PBS was mixed with 50 µl of alginate (final concentration: 10mg/kg alginic acid in PBS containing 10% DMSO). These three agents were administrated via intratumor injection biweekly starting 11 days post cancer cell injection. Tumor volume was measured once a week. All mice were sacrificed after 4 weeks of treatment.

### Abbreviations

| Abbreviation | Description |
| --- | --- |
| AI | Artificial Intelligence |
| BIAS | Biological Image Analysis Software |
| DVP | Deep Visual Proteomics |
| LGSC-Met | Low-Grade Serous Cancer - Metastasis |
| LGSC-PT | Low-Grade Serous Cancer - Primary Tumor |
| MS | Mass Spectrometry |
| PCA | Principal Component Analysis |
| ROIs | Regions of Interest |
| SBT | Serous Borderline Tumor |
| SBT-MP | Micropapillary Serous Borderline Tumor |

