## Supplementary Figures for "Spatial proteo-transcriptomic profiling reveals the molecular landscape of borderline ovarian tumors and their invasive progression"

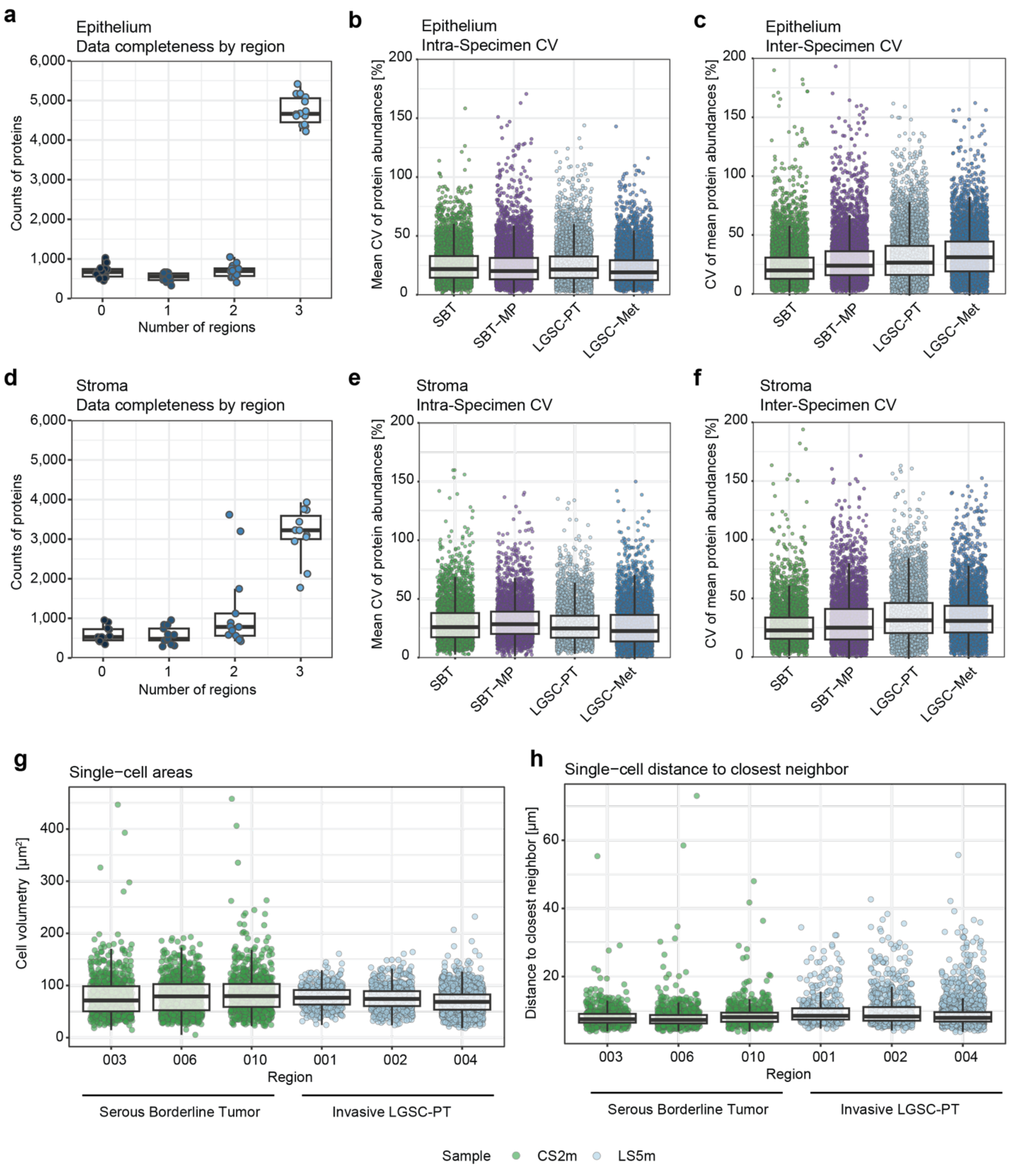


**Extended Data 1 | Deep Visual Proteomics - Quality control**

**a-f)** Deep Visual Proteomics (DVP) of the epithelial (a-c) and stromal (d-f) compartments. Protein counts in the epithelium (a) and the stroma (d), including all regions of interest from the same patient. More than 4,000 proteins were detected in all three selected regions of the same tissue specimen. Coefficient of variation (CV) for each of the cohort groups in the epithelium (b) and the stroma (e), showing data variability between regions within one slide. CV for each of the cohort groups in the epithelium (c) and the stroma (f) represent data variability between patients in the 4 different histologic categories.

**g-h)** Characteristics of Artificial Intelligence (AI) detected cells for DVP experiments shown by an exemplary patient in this study. DVP preserves spatial and morphological information of each extracted cell, enabling the depiction of cell size (g) and the distance to the closest neighboring cell (h), exemplarily shown for serous borderline tumor (SBT) and low-grade serous cancer (LGSC-PT). The cell size in LGSC is more uniform than in SBT, indicative of cancer cells (g), while the distance to the closest neighbor is higher, indicating the invasion of LGSC into the stroma (h).

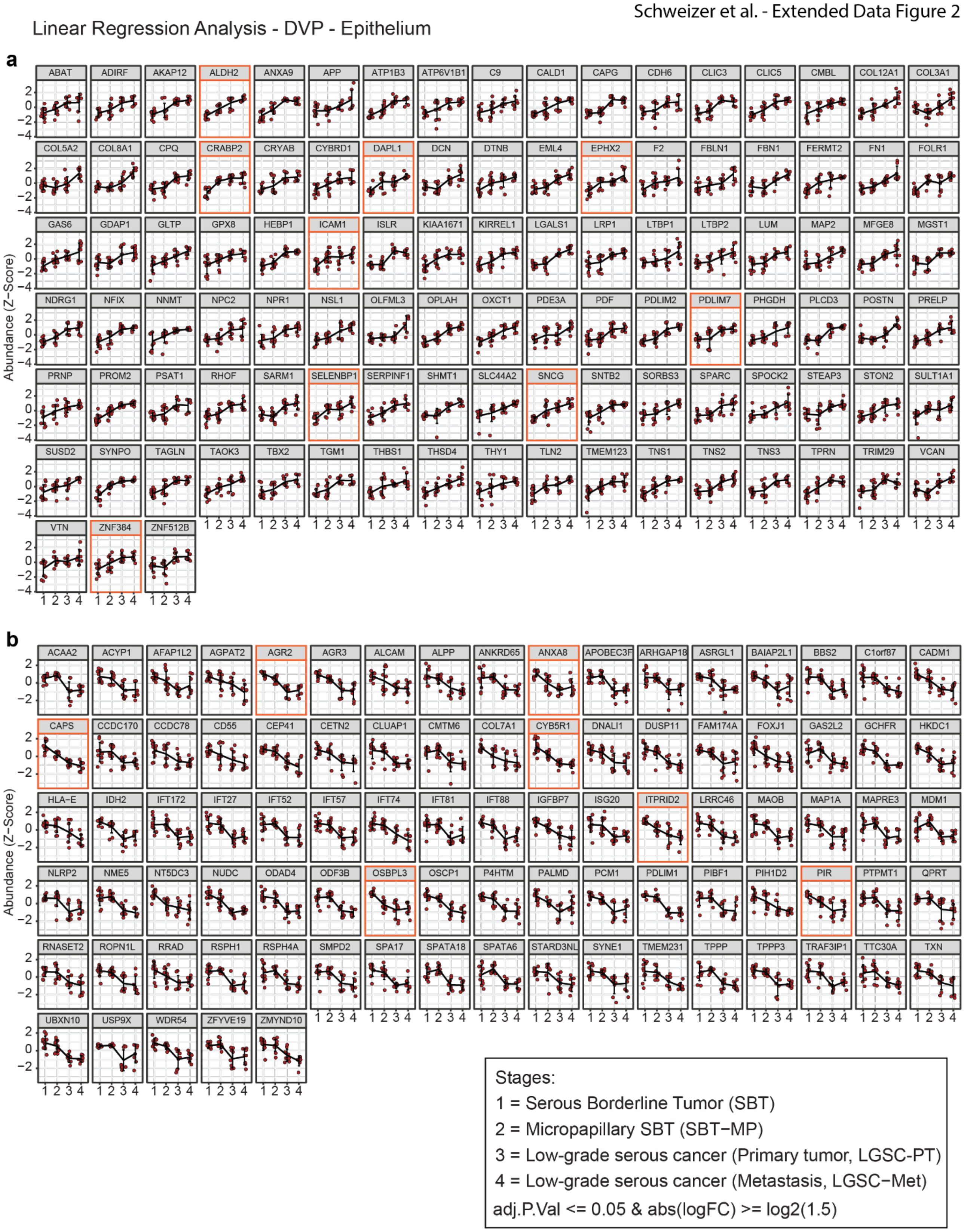


**Extended Data 2 | DVP - Linear regression analysis of changes in the epithelium from borderline tumors to metastatic low-grade serous cancer**

**a-b)** Proteins showing a linear increase (a) or decrease (b) in the epithelium. c) SBT-MP subgroup analysis. Boxplots representing proteins that show substantial increase from SBT to SBT-MP and persist or increase in abundance in LGSC and LGSC-Met. These proteins potentially characterize the progression from SBT- to SBT-MP, driving transformation as they persist in the invasive LGSC and its corresponding metastasis. Proteins showing substantial abundance changes between SBT and SBT-MP are highlighted by an orange frame. Results of the linear regression analysis after filtering for significant changes (adj. p -value <= 0.05, fold change > 1.5) are shown as line plots for each significant protein in the epithelial compartment (see also Supplementary Table 4).

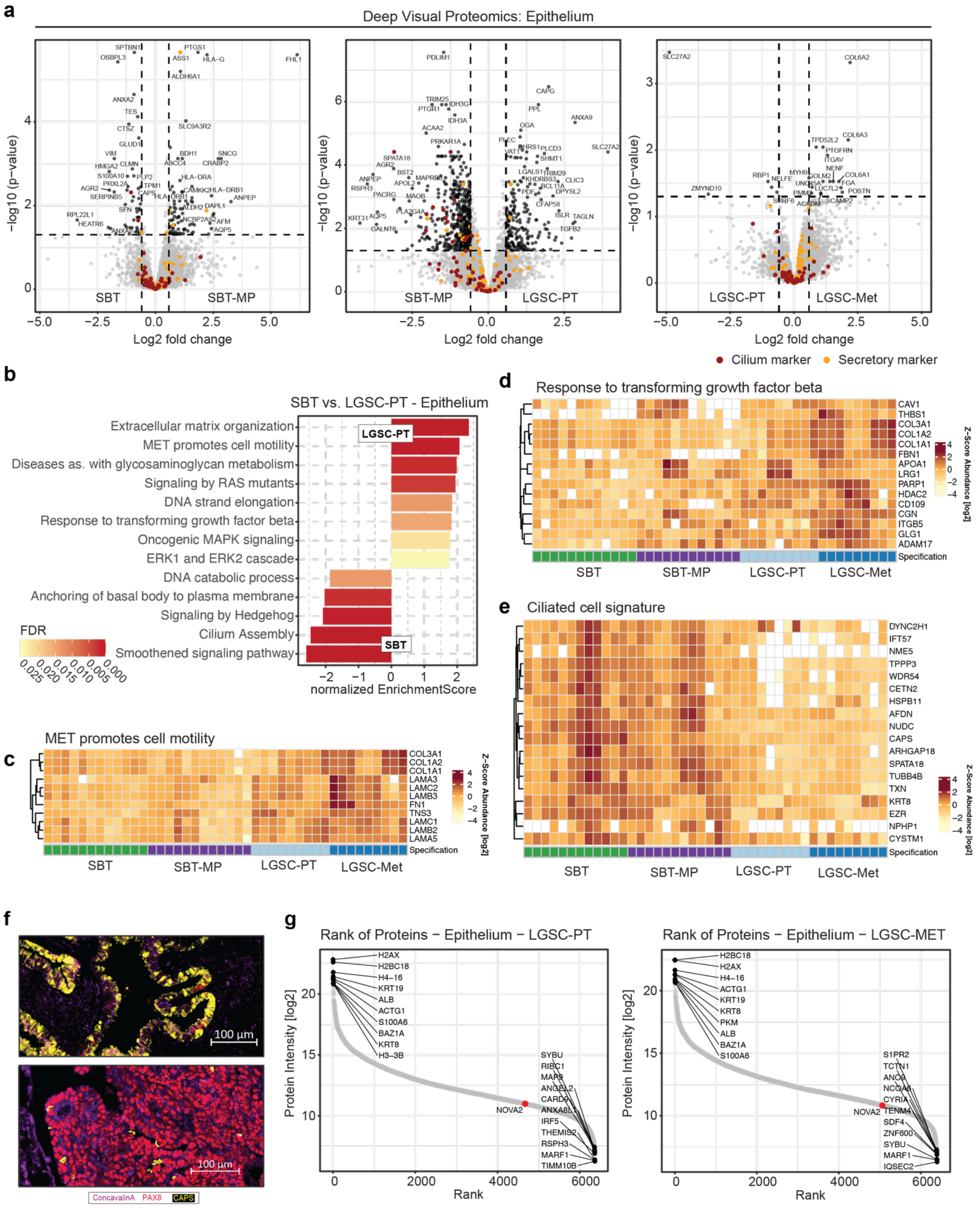


**Extended Data 3 | Comparison of epithelial protein expression during the progression of borderline tumors to metastatic low-grade cancer – proteomic analysis**

**a)** Volcano plots. Proteomic analysis of serous borderline tumors (SBT), micropapillary SBT, low-grade serous cancer (LGSC-PT), and corresponding metastasis (LGSC-Met) in the epithelial compartment (see regression analysis epithelium, Extended Data 5, for details). Proteins matching the significance for differential regulation (DR) criteria are highlighted in black, markers of secretory cells in orange and ciliated cells in red.

**b)** GSEA biological pathway enrichment comparing serous borderline and low-grade serous cancer. Selected terms are shown.

**c-d)** Heatmap, DVP. Proteins associated with the enriched biological terms (Pathway Reactome) 'MET promotes cell motility' (c), and 'Response to transforming growth factor β' (d) show an increase in abundance starting from SBT-MP to LGSC-Met.

**e)** Heatmap, DVP. Proteins enriched in ciliated cells showed a gradual decrease in protein intensity during the progression of the four histologies.

**f)** Immunofluorescence for PAX8 (red) and CAPS (yellow) stained in borderline and low-grade serous cancer metastasis confirming the lower proportion of ciliated cells in LGSC-Met compared to SBTs. Cell membranes were stained using ConcavalinA (purple).

**g)** Protein rank plots for LGSC-PT and LGSC-Met in the epithelial compartment. Proteins were ranked by MS intensity, and the highest and lowest ranking proteins were highlighted. Of note, NOVA2 is within the experimental detection limit but was not expressed in SBT and SBT-MP (Fig. 2f-h).

Serous borderline tumor (SBT), micropapillary SBT (MP-SBT), low-grade serous tumor primary tumor (LGSC-PT), and corresponding metastases (LGSC-Met).

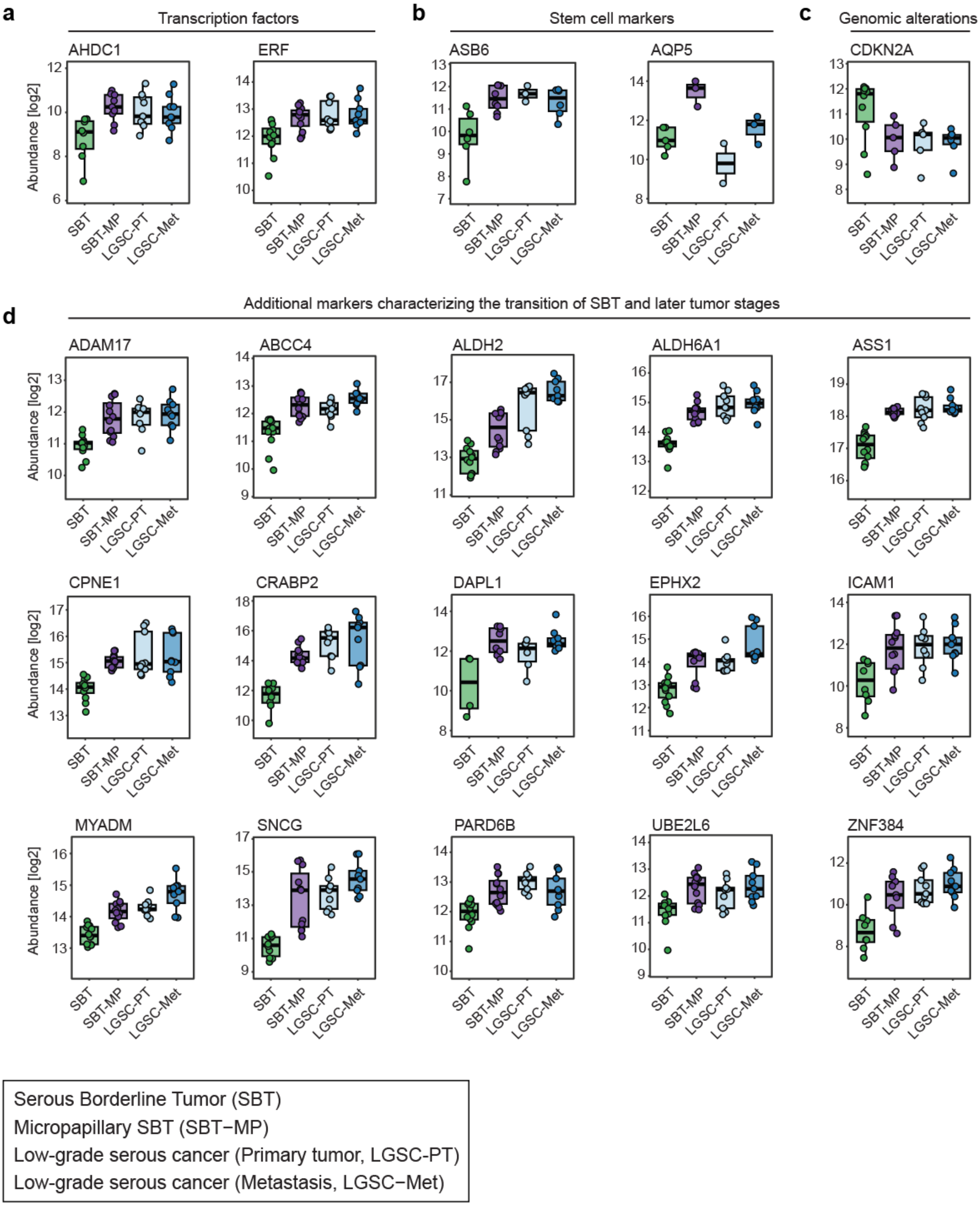


**Extended Data 4 | DVP – Early changes of epithelial protein expression in the transition to invasive phenotypes**

Proteins elevated in SBT-MP which largely maintain expression in LGSC-PT and LGSC-Met thereafter. Transcription factors (a), stem cell markers (b) and CDKN2A altered on the genomic level (c) are specifically annotated. Additional markers characterize the early transition of SBT to more invasive phenotypes (d). Boxplots show clear differences between SBT and the other groups. Of note, the y axis has different scales for every protein and does not start at 0.

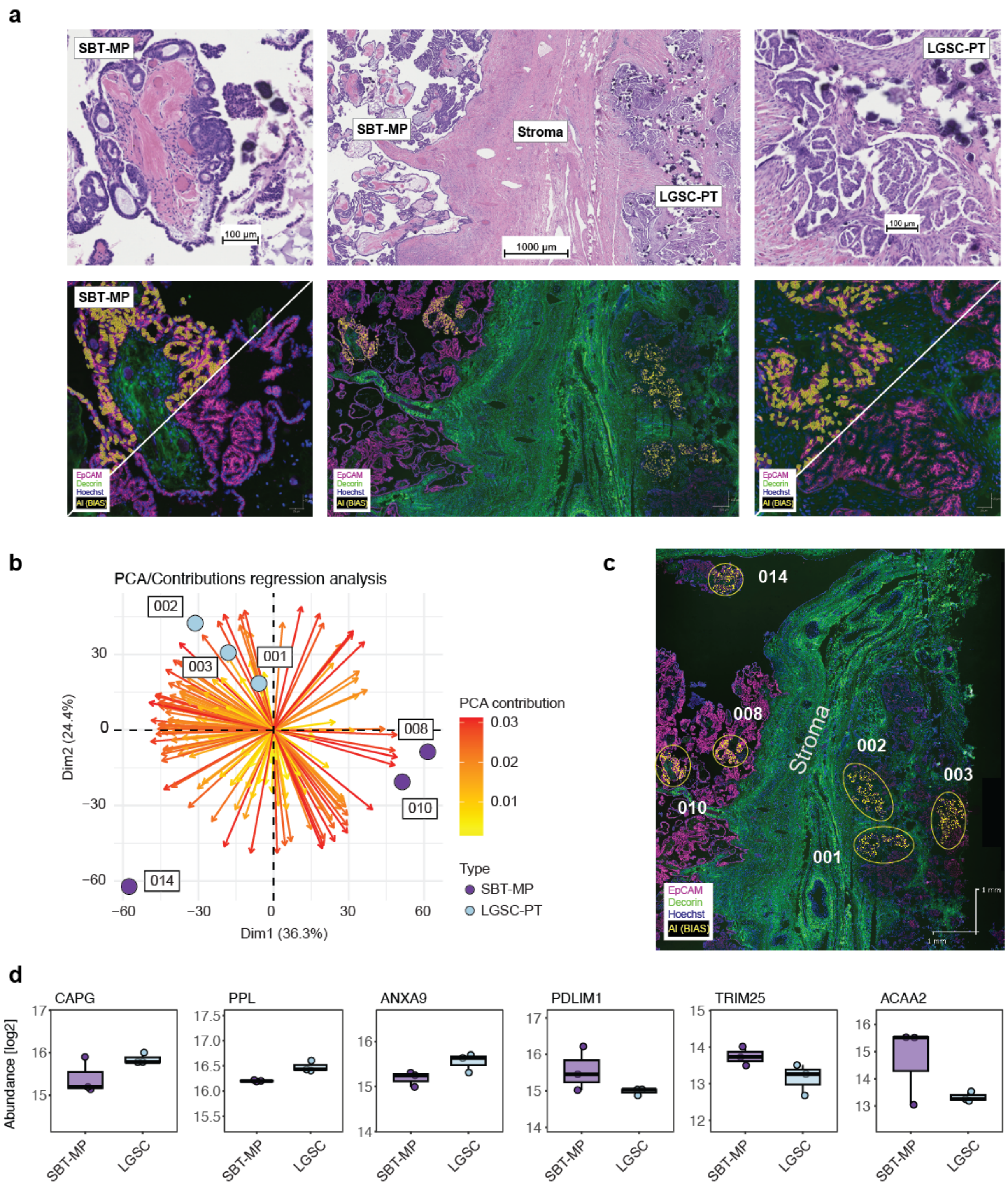


**Extended Data 5 | Detailed analysis of an invasive low grade serous cancer adjacent to a serous borderline tumor with micropapillary features.**

**a)** Case study of a woman with a serous borderline tumor with micropapillary features and low-grade serous cancer. H&E staining (upper panel) and AI-based cell recognition using immunofluorescence (EpCAM-purple, decorin-green) below the white diagonal line, followed by AI segmentation (yellow).

**b)** Principal Component Analysis for the epithelium separating the two histologies in both dimensions. The PCA is shown as a biplot of individual regions (points) and proteins (arrows) representing proteins of significant linear increase or decrease (Extended Data Fig. 3) across the investigated transition. Of note, region 8/9 (LMP) are the closest to LGS region 1-3, but region 014 was much farther away and did not cluster with the other samples of SBT-MP.

**c)** Immunofluorescence images (a) showing, on one slide, LMP (region 008, 010, 014) and LGSC (region 001, 002, 003). Of note, region 014 is further away from the tumor-stroma interface, consistent with the PCA (b).

**d)** Exemplary boxplots for the previously identified proteins (Extended Data Fig. 2b) for the transition from SBT-MP to LGSC.

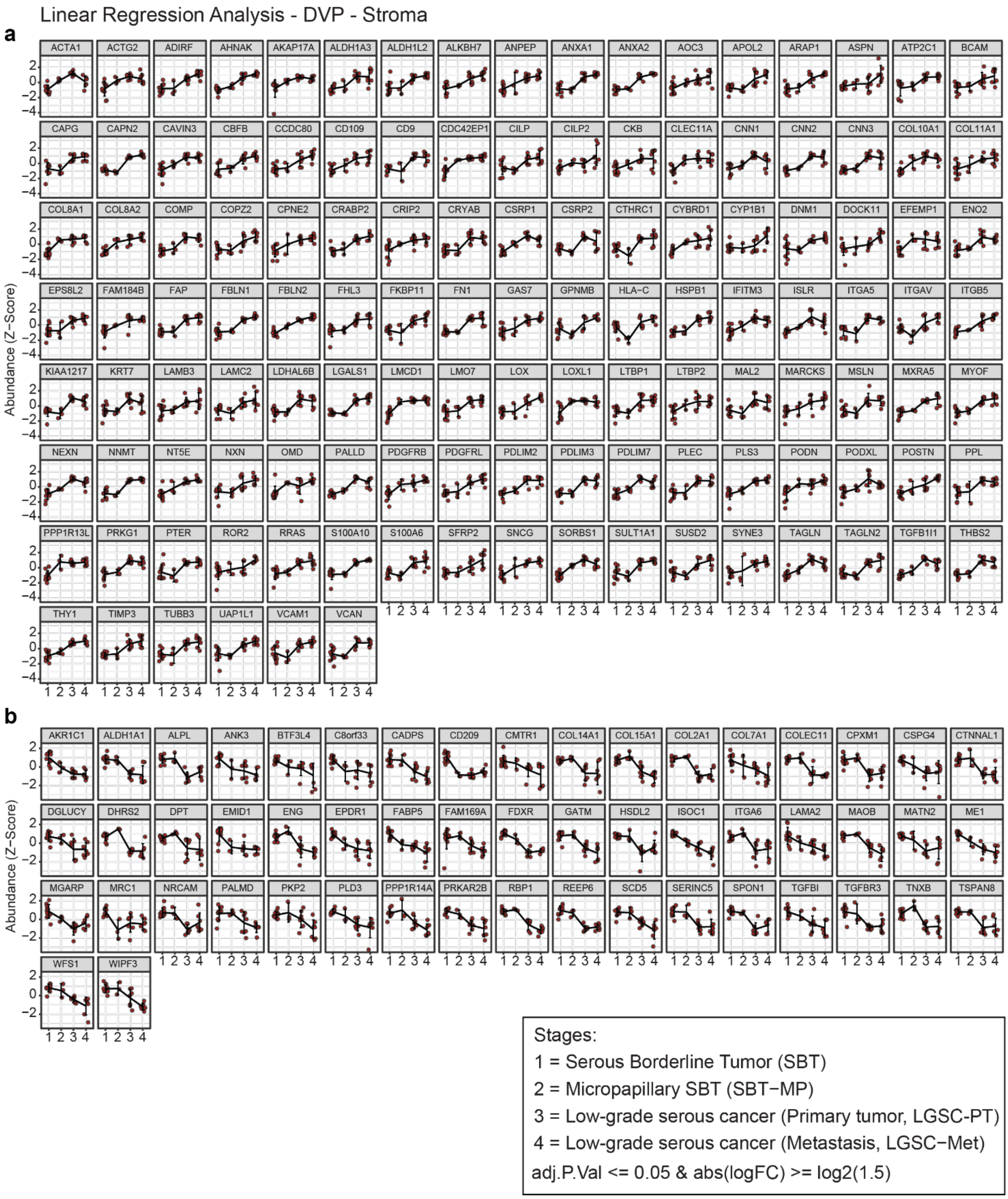


**Extended Data 6 | DVP - Linear regression analysis of protein changes in the stroma from borderline tumors to metastatic low-grade serous cancer**

**a-b)** Stromal proteins showing a linear increase (a) or decrease (b) in the 4 cohorts. Results of the linear regression analysis after filtering for significant changes (significance threshold: adj. p -value <= 0.05, log fold change > 1.5) in protein expression are shown as line plots for each significant protein in the stroma (see also Supplementary Table 4).

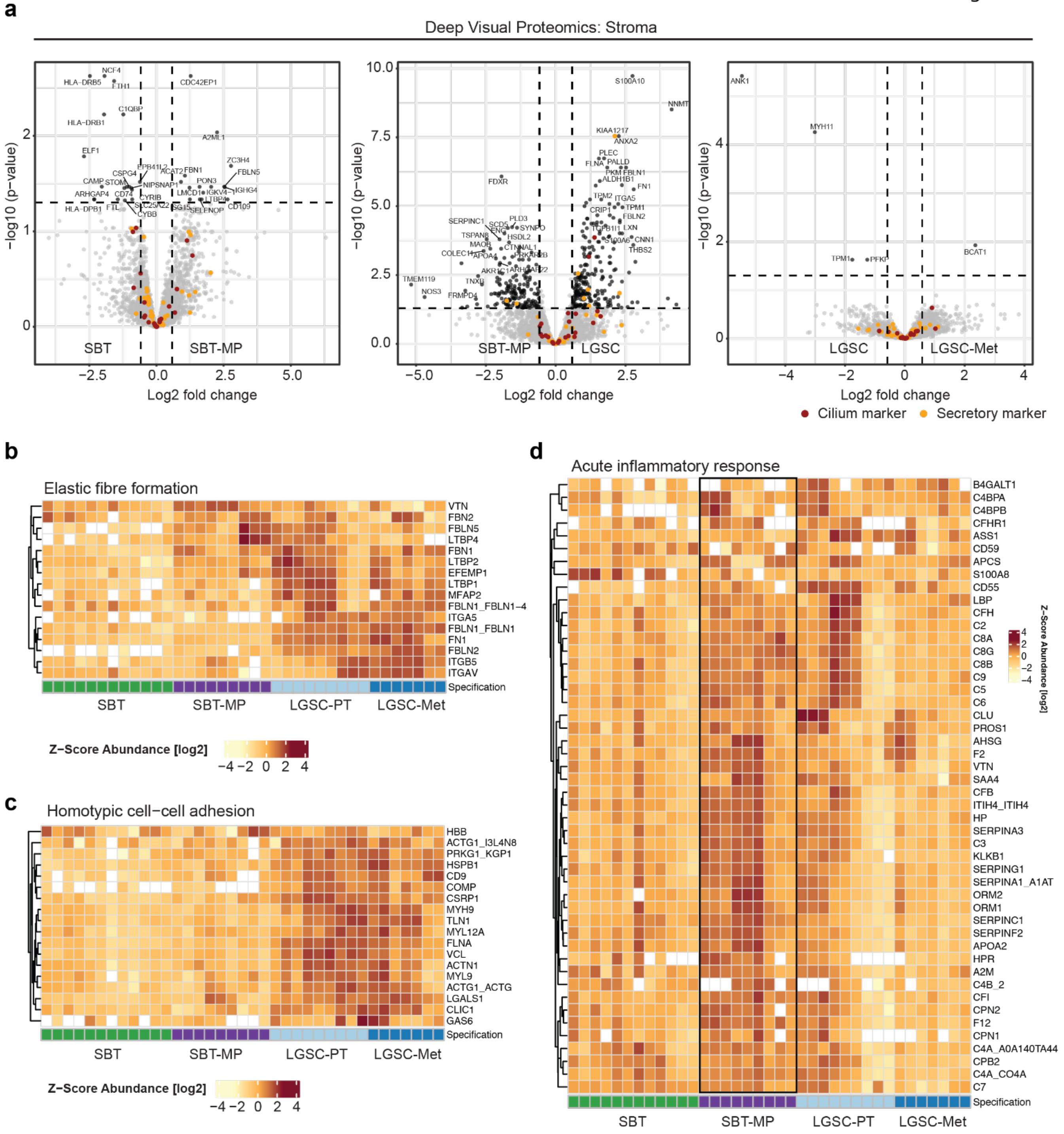


**Extended Data 7 | Proteomic analysis of changes in the stroma from borderline tumors to metastatic low-grade serous cancer using Deep Visual Proteomics**

**a)** Volcano plots. Stroma. Proteomic analysis of borderline tumors, micropapillary borderline, low-grade serous cancer, and corresponding metastasis in the stromal compartment.

**b-d)** Heatmap. Stroma. Proteins involved in elastic fiber formation (b) and cell-cell adhesion (c) show increased abundance from SBT to LGSC-Met, while there was an increase in inflammation in the stroma of micropapillary tumors (d).

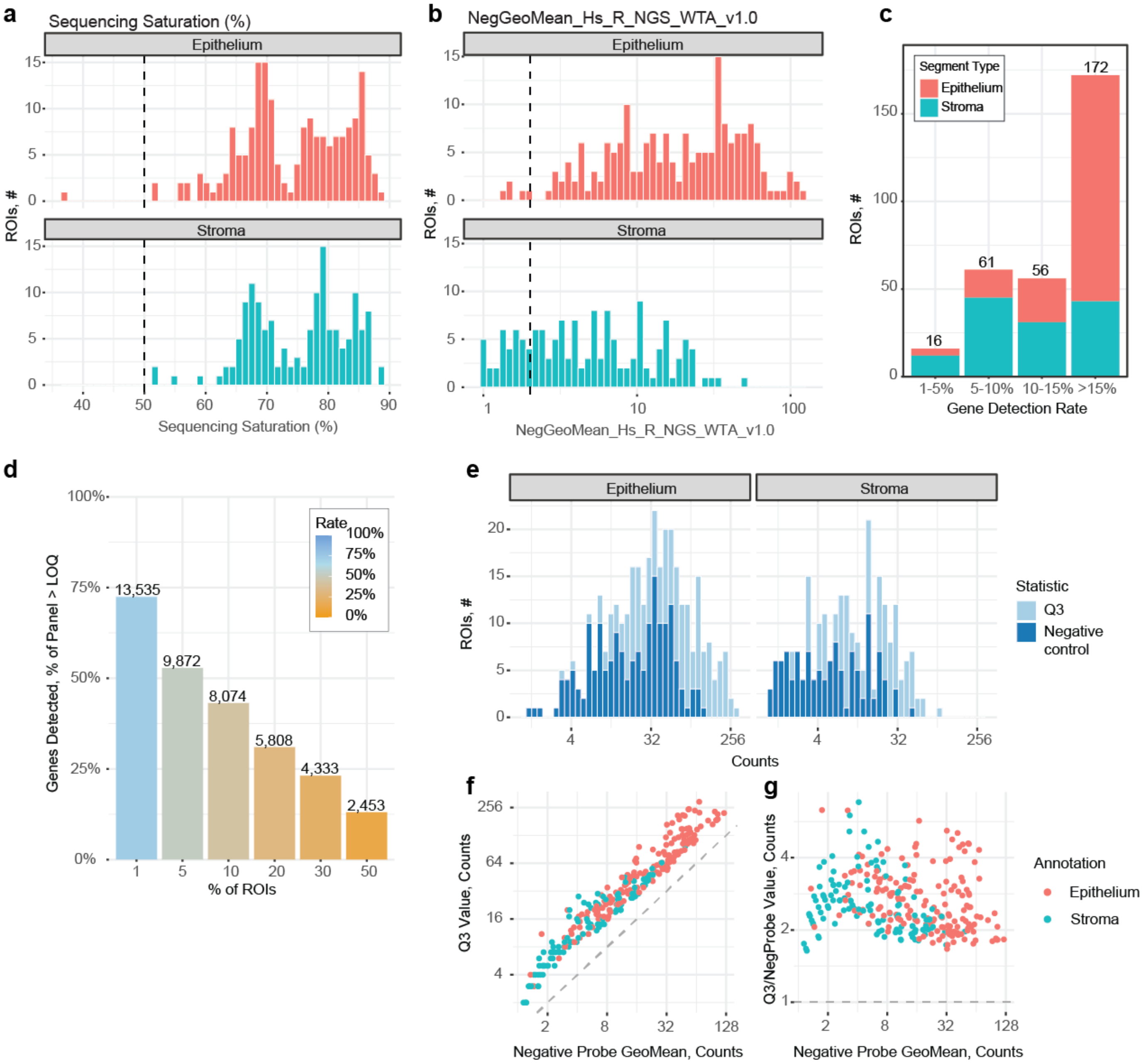


### Extended Data 8 | Spatial transcriptomics – Quality control

**a)** Distribution of the sequencing saturation in epithelium and stroma, respectively. A high sequencing saturation indicates the reproducible alignment of reads-to-target sequences. Quality control (QC) filtered samples are left of the dashed line.

**b)** Negative geometric means based on negative control samples for each compartment. QC filtered samples are located left of the dashed line.

**c)** Gene detection rate (detected genes/ genes in whole transcriptome atlas (WTA)) in regions of interest (ROIs) of the investigated compartments. ROIs < 5% gene detection rate were excluded from the analysis. The Nanostring whole transcriptome atlas > 18,000 genes.

**d)**Detected genes in different percentages of ROIs. Genes detected in less than 5% of ROIs were excluded from the analysis.

**e-g)** Comparison of the upper quartile (Q3) of counts in each ROI compared to the geometric mean of the negative control probes. A good separation of the Q3 and negative probe counts at both the distributions (e) and per segment levels (f, g) validates the quality of the previous filtering process.

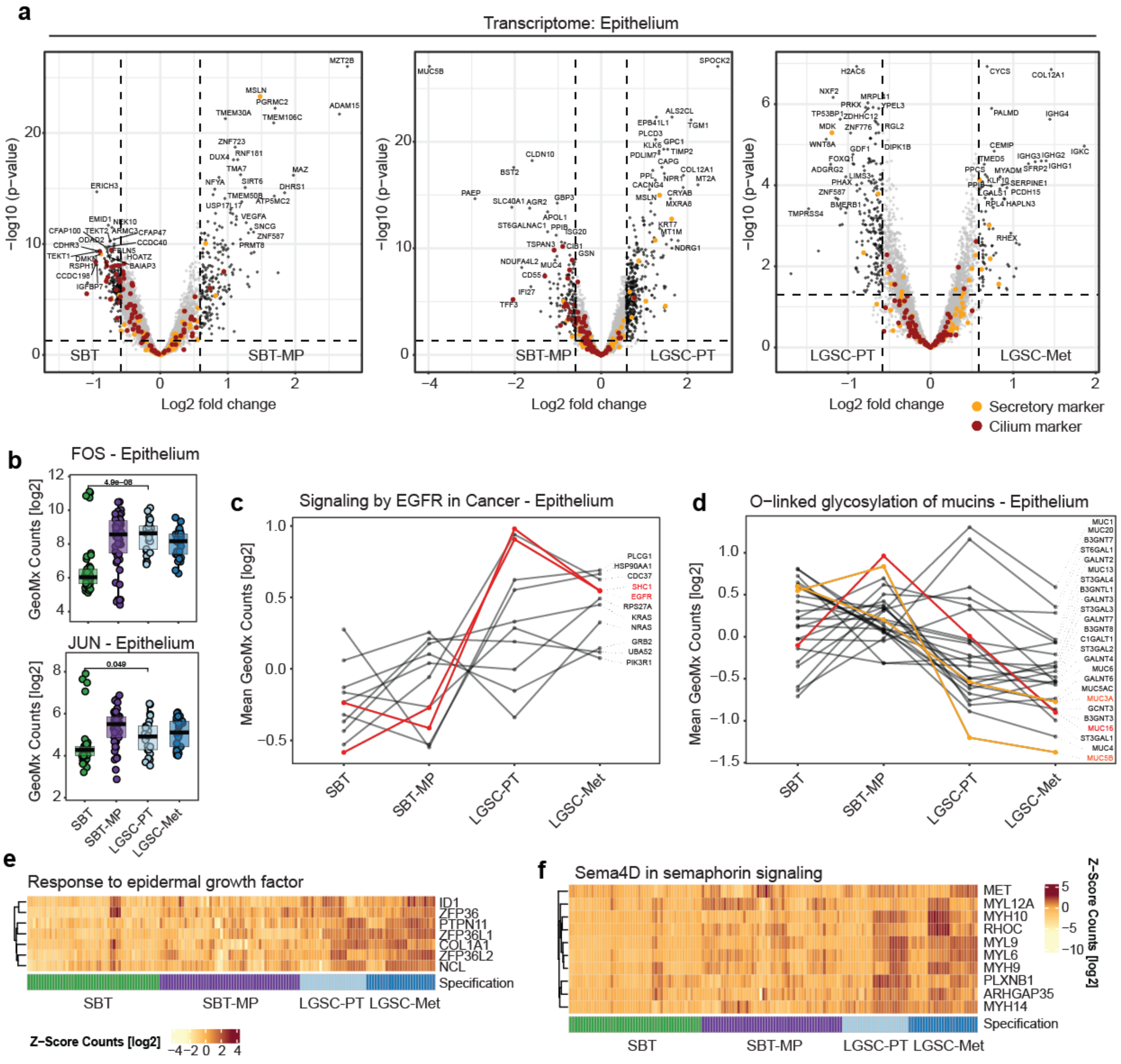


**Extended Data 9 | Spatial transcriptomics study of changes in gene expression during the progression of borderline tumors to metastatic low-grade cancer in the epithelium**

**a)** Volcano plot. Epithelium. Nanostring analysis of the transition from SBT to invasive LGSC-Met. Transcripts for ciliated and secretory cells are highlighted in orange and red, respectively.

**b)** Boxplots showing the GeoMx counts for the transcription factors JUN and FOS across the progression series (Student's t-test).

**c-d)** Profile plots of pathway-associated transcripts depicted in (Fig. 4d) for 'EGFR signaling' (c) and 'O-linked glycosylation of mucins' (d).

**e-f)** Heatmap, Transcriptome. Transcripts associated with the enriched biological terms 'response to epidermal growth factor' (e) and 'Sema4D in semaphoring signaling' (f) show an increase starting from SBT-MP to LGSC-Met.

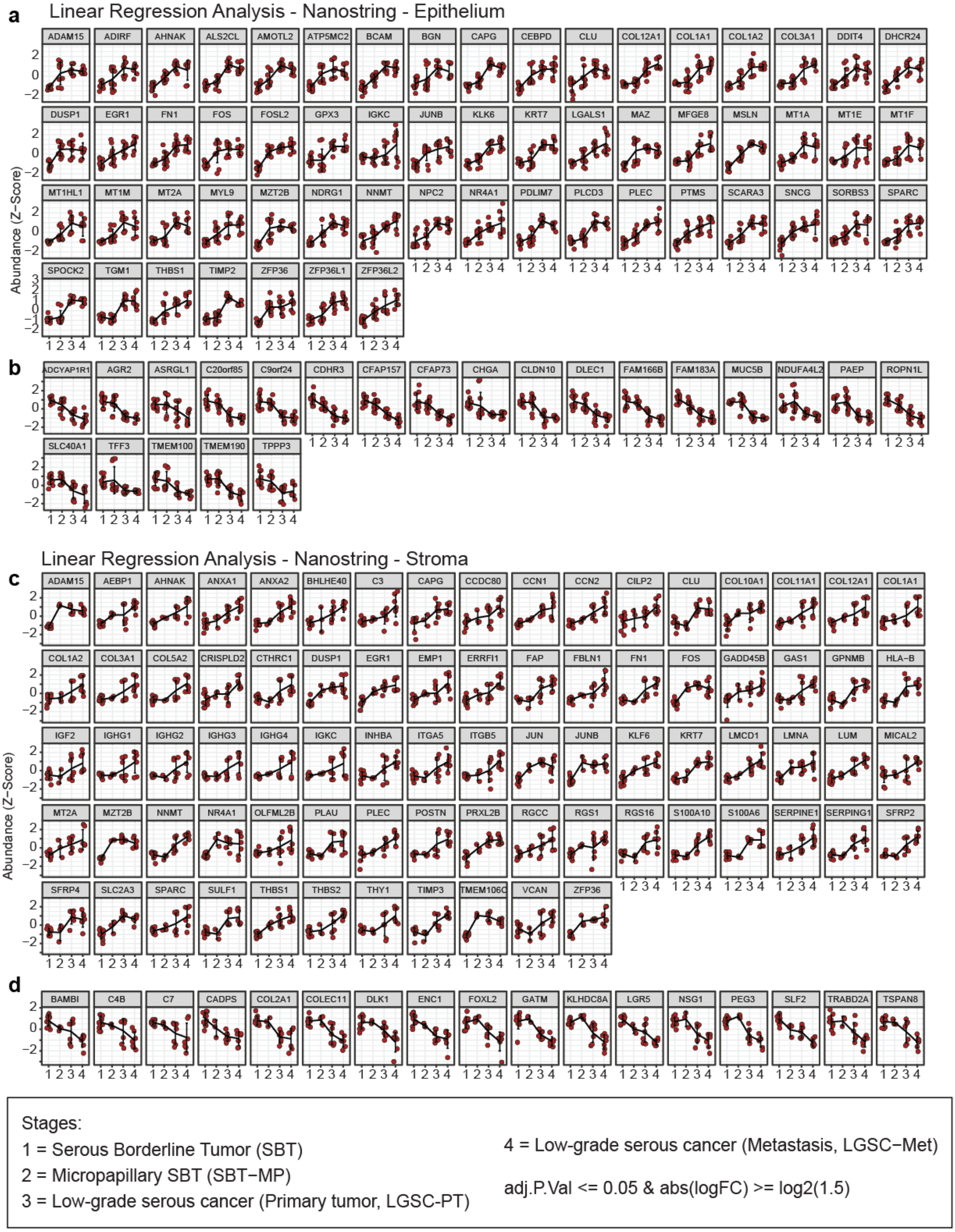


**Extended Data 10 | Spatial Transcriptomics - Linear Regression of changes in gene expression in the epithelial and stromal compartment from borderline tumors to metastatic low-grade serous cancer**

**a)** Transcripts showing a linear increase (a) or decrease (b) in the epithelium.

**b)** Transcripts showing a linear increase (c) or decrease (d) in the stroma.

Results of the linear regression analysis after filtering for significant changes (adj. p -value ≤ 0.05, fold change > 1.5) shown as line plots for each significantly changed gene transcript in the epithelium (a) and stroma (b) (see also Supplementary Table 4).

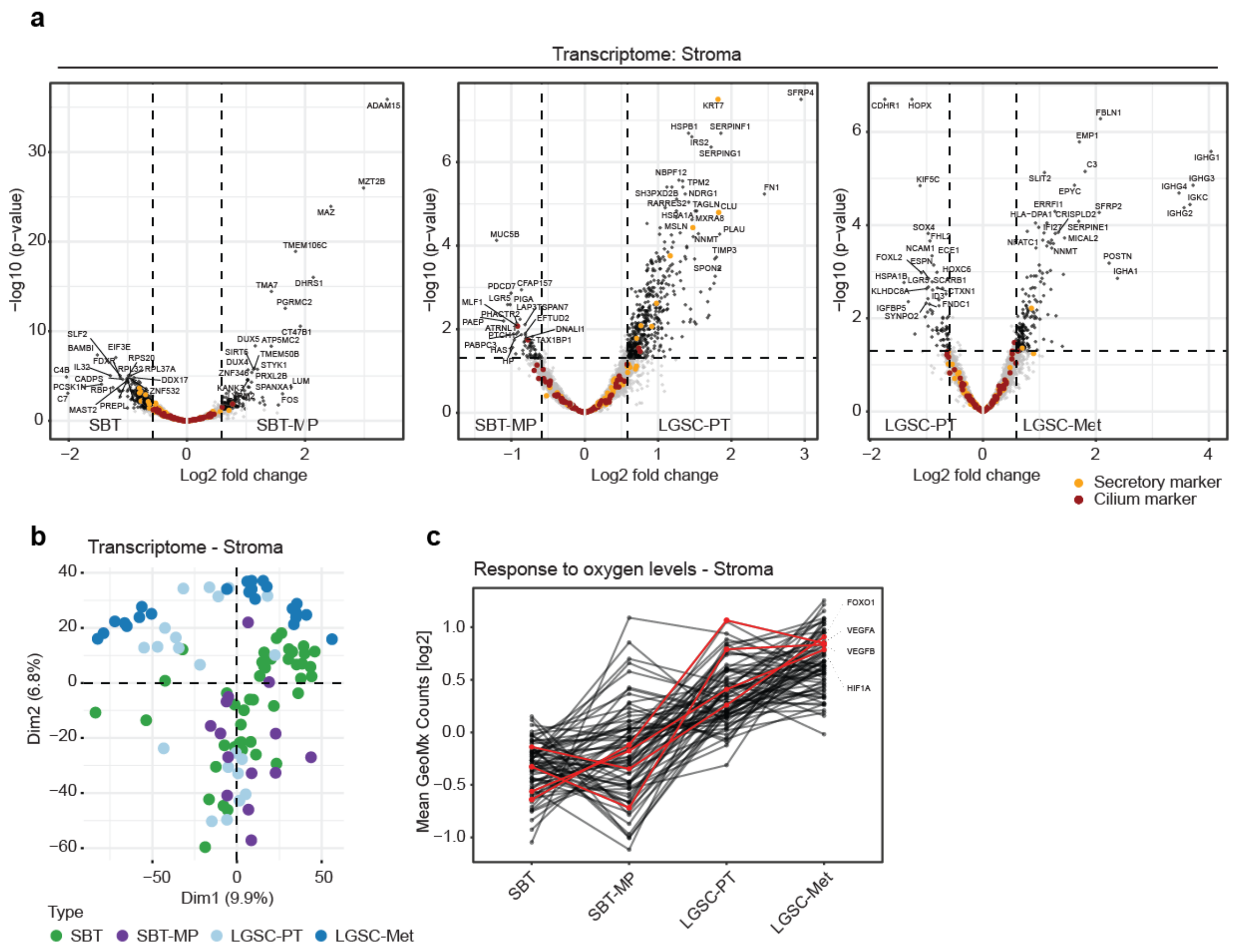


### Extended Data 11 | Spatial transcriptomics analysis of the progression of borderline tumors to metastatic low-grade cancer in the stroma

**a)** Volcano plot. Stroma. Nanostring analysis of the transition from SBT to invasive LGSC-Met. Transcripts for ciliated and secretory cells are highlighted in orange and red, respectively.

**b)** Nanostring Principal Component Analysis for transcripts detected in the stroma for the indicated histologies.

**c)** Profile plots of pathway-associated transcripts shown in Fig 4j for the ‘Response to oxygen levels’. Proteins with critical roles in the pathway (FOXO1, VEGFA, VEGFB, HIF1A) are annotated in red. All other transcripts are summarized in Supplementary Table 6.

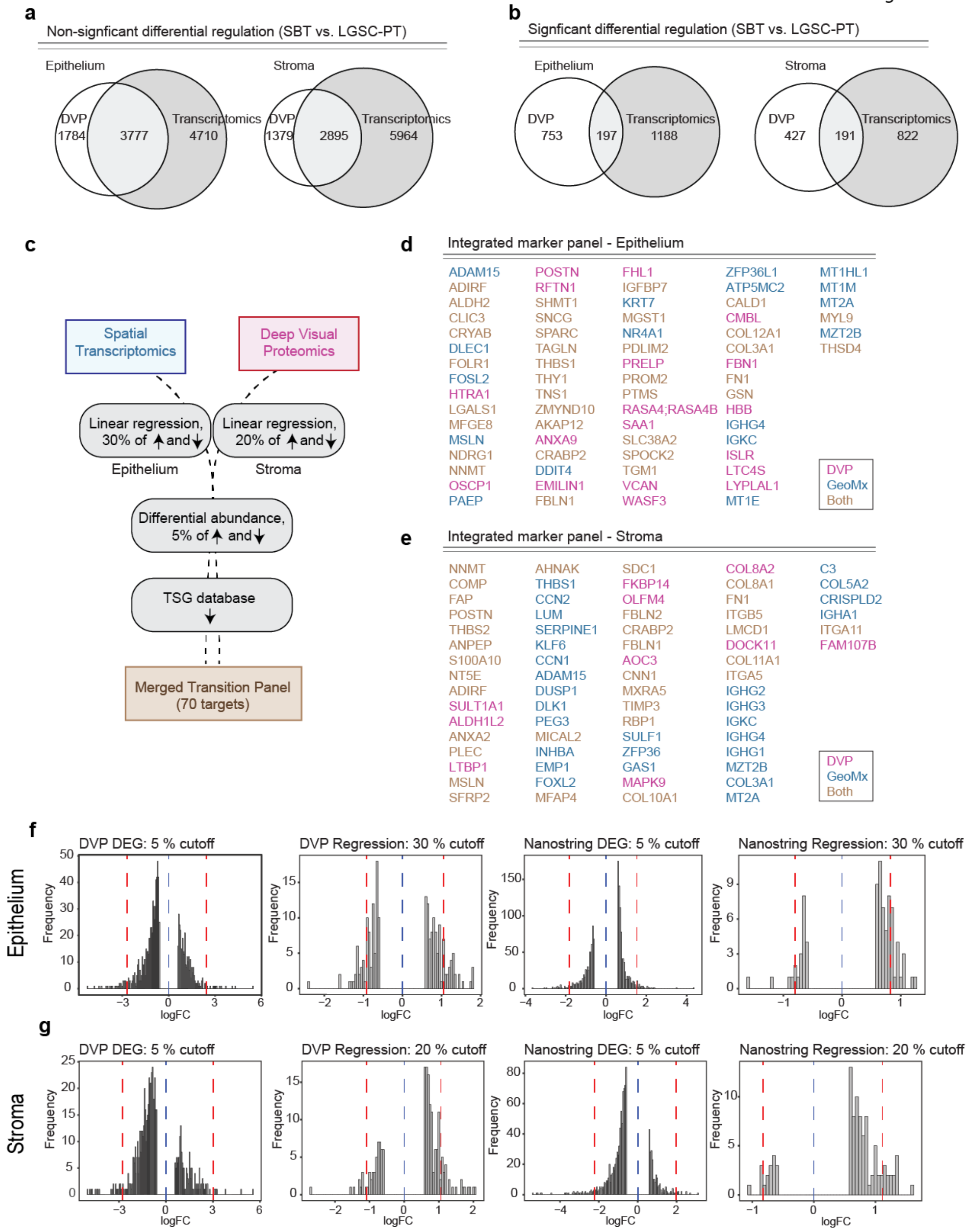


**Extended Data 12 | Integration of spatial proteomics and transcriptomics generating an annotated list of putative progression markers**

**a)** Venn diagrams illustrating proteins/transcript without significant differences between SBT and LGSC-PT for epithelium and stroma

**b)** A similar analysis was performed for the overlap and unique findings of significantly differentially regulated proteins/transcripts for both technologies, comparing SBT and LGSC-PT for epithelium and stroma.

**c)** Integration of protein/transcripts with potential biological relevance in the transition from SBT to LGSC-PT. A final list of substantial changes between SBT and LGSC-PT in either compartment was generated in three steps. First, the 30% and 20% most substantially altered proteins/transcripts (by fold change) were selected from the regression analysis in the epithelium and stroma, respectively, in both omics datasets (Supplementary Table 4). Second, the top 5% differentially expressed proteins/transcripts (by fold change) in the epithelium and stroma in the comparison of LGSC and SBT were selected from both omics datasets, while excluding targets identified in the first step (Supplementary Table 5, 9). Third, the subset of downregulated genes/proteins was annotated and filtered to select for known tumor suppressors using the 'Tumor suppressor gene (TSG) database'.

**d, e)** 70 most relevant proteins/transcripts in both the epithelial (d) and stromal (e) compartments after multi-omic data integration (c) (Supplementary Table 10).

**f, g)** Histograms visualizing the cutoffs used to generate the tables in (d) and (e).

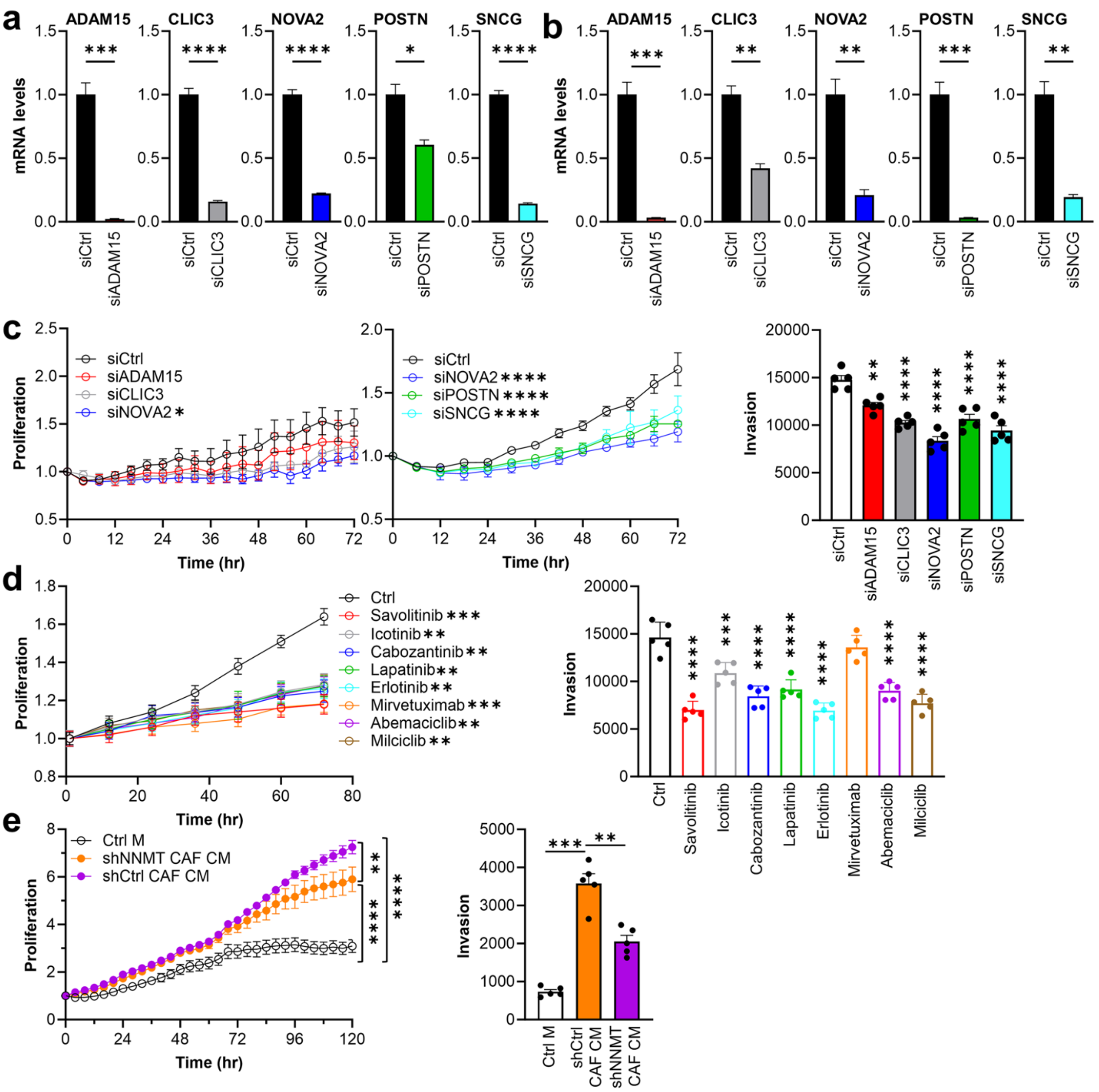

**Extended Data 13 | Functional studies characterizing the significantly changed 'omics' genes and pathways in the transitions of SBT to LGSC using LGSC cell lines.**

**(a,b)** Validation of positive siRNA screening results using gene knockdown via siRNA in the LGSC cell lines VOA4627 (a) and VOA6406 (b).

**c)** Validation of positive siRNA screening results in a second cell line, VOA6406, measuring proliferation (left) and invasion (right).

**d)** Functional testing of FDA approved inhibitors against the positive hits from the integrative "omics" approach (Extended Data 12) using the VOA6406 cell line for proliferation (left) and invasion (right) assays

**e)** Inhibition of NNMT in cancer associated fibroblasts (CAFs) reduces epithelial cell proliferation. VOA6406 LGSC epithelial cells were treated with either conditioned media from immortalized human CAFs where NNMT was inhibited using shRNA (shNNMT CAF CM), shRNA control transfected CAF condition media (shCtrl CAF CM), or control media (Ctrl M).

All growth curves and bar graphs show mean +/- SEM. The data presented in c-e was repeated in 3 independent experiments. Significance levels by p-value: * 0.05, ** 0.01, ***, 0.001, *** 0.0001.
